# Prevalence of multimorbidity combinations and their association with medical costs and poor health: a population-based study of U.S. adults

**DOI:** 10.1101/2022.01.18.22269483

**Authors:** Nicholas K. Schiltz

## Abstract

**BACKGROUND:** Multimorbidity is common, but the prevalence and burden of the specific combinations of coexisting disease has not been systematically examined in the general U.S. adult population.

**OBJECTIVE:** To identify and estimate the burden of highly prevalent combinations of chronic conditions that are treated among one million or more adults in the United States.

**METHODS:** Cross-sectional analysis of U.S. households in the Medical Expenditure Panel Survey (MEPS), 2016–2019, a large nationally-representative sample of the community-dwelling population. Association rule mining was used to identify the most common combinations of 20 chronic conditions that have high relevance, impact, and prevalence in primary care. The main measures and outcomes were annual treated prevalence, total medical expenditures, and perceived poor health. Logistic regression was used to calculate adjusted odds ratios and 95% confidence intervals.

**RESULTS:** Frequent pattern mining yielded 223 unique combinations of chronic disease, including 74 two-way (dyad), 115 three-way (triad), and 34 four-way combinations that are treated in one million or more U.S. adults. Hypertension-hyperlipidemia was the most common two-way combination occurring in 30.8 million adults. The combination of diabetes-arthritis-cardiovascular disease was associated with the highest median annual medical expenditures ($23,850, [Interquartile range: $11,593 – $44,616]), and the combination of diabetes-arthritis-asthma/COPD} had the highest age-race-sex adjusted odds ratio of poor self-rated health (adjusted odd ratio: 6.9, [95%CI: 5.4 – 8.8]).

**CONCLUSION:** This study demonstrates that many multimorbidity combinations are highly prevalent among U.S. adults, yet most research and practice-guidelines remain single disease focused. Highly prevalent and burdensome multimorbidity combinations could be prioritized for evidence-based research on optimal prevention and treatment strategies.

## INTRODUCTION

An estimated 42% percent of American adults 18 and older suffer from multiple chronic conditions (i.e. multimorbidity).^1^ Persons with multimorbidity have higher mortality rates, lower health-related quality life, increased healthcare use, and are at higher risk for many poor health outcome events including adverse drug events.^2–6^

Despite the high frequency of multimorbidity in the population, most clinical practice guidelines, and the evidence they are based upon are single disease focused. Adhering to multiple, sometimes conflicting, guidelines for multiple individual diseases is burdensome^7^ and conveys considerable risk to the patient in terms of drug-drug or drug-disease interactions.^8,9^ The overall optimal treatment plan for a given patient with multimorbidity is unlikely to be the linear combination of the best treatment for each individual condition.^9,10^ There is a clear need to develop guidelines, care management plans, and patient education programs that meet the needs of patients with multimorbidity. Understanding which conditions most commonly coexist together and their impact on outcomes is a critical first step to establish research priorities for building the evidence-base necessary to achieve this.

Understanding the rate that disease coexist or cluster together at higher-than-expected rates, may also provide clues about the etiology between two or more conditions. For example, inflammation has been shown to be a mechanism for numerous conditions including cancer, cardiovascular disease, chronic kidney disease, and diabetes.^11^ Damage caused by vascular diseases may be a cause of Alzheimer’s Disease and other dementias.^12^ Understanding these shared risk factors and pathways could lead to the development of medications or behavioral interventions that target more than one condition simultaneously, hence reducing polypharmacy and treatment burden in this population.

While numerous data exist on the prevalence of single chronic conditions^1,13^, much less is known about the prevalence of *specific combinations* of multimorbidity. Most prevalence studies of multimorbidity are based on counts of conditions.^14,15^ However, measuring the impact of multimorbidity based on counts of conditions or weighted scores may miss important heterogeneity, because the specific combinations of chronic disease matter in terms of health outcomes^16^ including mortality and poor health,^5^ activity of daily living and instrumental activity of daily living limitations,^17,18^ cost and utilization,^6^ and quality of care.^19^

Studies that have examined the prevalence of specific combinations of multimorbidity in the US have been mostly limited to dyad and triad combinations,^20–23^ a small number of conditions,^17,24^ conducted in a specific sub-populations^21^ or geographic areas,^25^ or limited to older adults.^24,26^ The prevalence of specific multimorbidity combinations of the general community-dwelling adult population has not been established comprehensively in the United States.

The goal of this study was to fill this knowledge gap and identify and rank the most frequent multiway (two-way, three-way, four-way, etc…) combinations of multimorbidity treated among adults in the United States. An innovative machine learning algorithm –association rule mining – was used to identify all combinations of multimorbidity occurring in one million or more U.S. adults. The association with average and median per capita direct medical expenditures and perceived fair or poor health status was also examined, to identify combinations associated with the most burden.

## METHODS

### Design, Setting, and Participants

This study was a cross-sectional analysis of the 2016 – 2019 Medical Expenditure Panel Survey Household Component (MEPS). The MEPS is a large-scale panel survey of a nationally representative sample of households in the United States and is administered by the Agency for Healthcare Research and Quality (AHRQ).^27^ The MEPS collects data at the person-level on numerous topics including medical expenditures, utilization, medical conditions, health status, and demographics. Interviews are conducted in-person using computer-assisted personal interview (CAPI) technology. The MEPS is designed in a way that it can be used for both longitudinal analysis over a two-year period, and cross-sectional analysis to obtain annual estimates in a single year,^28^ or pooled across multiple years.^29^ This study uses cross-sectional methods and weights as the main goal was to produce national estimates of prevalence. The 2016 – 2019 data was chosen as these are the most recent currently available data and earlier years used ICD-9-CM coding for medical conditions. Multiple years of data were pooled together to provide a larger sample size as some combinations will occur in less than 1% of the total study population.^29^ This study included all adults age 18 and older with non-zero weights. Subjects with missing data on self-reported health or expenditures were excluded (n≤10). The final unweighted sample was 89,947 adults (Supplementary Figure 1).

### Ethics Statement

The MEPS data used in this study is publicly available and de-identified. The Case Western Reserve University Institutional Review Board reviewed the protocol for this study and deemed it to be exempt under U.S. federal law.

### Measures

Health conditions were self-reported through open-ended questions about conditions reported to be treated with prescription medications or associated with health care utilization.^30^ The respondents verbatim text responses were coded according to the International Classification of Diseases, Tenth Revision, Clinical Modification (ICD-10-CM). Treatment of these conditions may have been received through inpatient stays, outpatient or office-based visits, emergency department visits, home health care, or prescribed medications. Chronic conditions were then classified into one of 20 chronic conditions used in recent studies of multimorbidity, because of their high relevance, impact, and prevalence among primary care clientele.^31–33^ A list of each chronic condition categories and corresponding ICD-10-CM codes is included in the appendix (Supplementary Table 1).

Perceived health was assessed by asking respondents to rate their health as excellent (1), very good (2), good (3), fair (4), or poor (5). The variable was dichotomized into two values: a response of excellent, very good, or good was categorized as good perceived health, while a response of fair or poor was categorized as poor perceived health.

Medical expenditures were defined as the sum of direct payments for care provided during the year, including out-of-pocket payments and payments by private insurance, Medicaid, Medicare, and other sources. Expenditure data were collected in the MEPS through both self-report and through data collected from the respondents’ physicians, hospitals, and pharmacies, when available. The MEPS provides imputed values when data are missing or payments are made under capitated plans.^34^ Total expenditures were adjusted for annual inflation and are reported in terms of 2019 dollars using the Personal Consumption Expenditure Health Price Index from the Bureau of Economic Analysis.^35^ Out-of-pocket expenditures were adjusted for inflation using the Consumer Price indices for medical care (CPI–M).^35^

### Statistical Analysis

The main analytic techniques in this study were association rule mining and frequent pattern mining.^36^ In brief, association rule mining can be thought of as a two-step process. In the first step (frequent pattern mining) all combinations of items with a minimum support (i.e. prevalence in the study population) are discovered. In the second step, “association rules” of the form X = > Y are created, where X is one or more items (chronic conditions in this study) and Y is a single-item consequent (poor health or medical expenditures in our study) that X is associated with.

The method was applied here by treating each subject in the study data as the “transaction,” and each chronic condition as the “items” to find the most frequently coexisting combinations out of the 1,048,576 (2^20^) possible combinations of 20 conditions. The minimum support (prevalence) threshold was set at 0.20% for the initial pass, and then after applying survey weights, any combinations with a point estimate of less than one million adults (∼0.40% of the U.S. adult population) was dropped.

The MEPS uses a complex stratified random sampling design. Design-based survey methods (i.e. Taylor series estimation) were used to account for stratification and clustering effects when estimating variances.^37^ Person-level weights in each year were divided by four (number of total years of data) and applied to get the annualized estimated number and percent of persons with a disease combination and 95% confidence interval.^28^ Likewise, the median and average annual per capita medical expenditures, per capita out-of-pocket expenses, and percent self-reporting poor health status was calculated for each combination after applying weights.

Combinations of conditions and corresponding prevalence estimates represent “at least”, rather than “exactly.” For example, people with hypertension-hyperlipidemia-diabetes would be a subset of the people with hypertension-diabetes.

For each combination, we calculated the observed-to-expected prevalence ratio (also known as lift), which is the observed prevalence of the combination divided by the expected prevalence given all single diseases in the combination are independent of each other. Lift significantly greater (or lower) than 1 indicates the coexistence of diseases is unlikely due to chance alone.

To identify specific combinations associated with the highest average cost and perceived poor health burden, we filtered out combinations that were redundant in that they offered little new information on the outcome measure over a more parsimonious (i.e. superset) combination, using a minimum improvement criteria of 10%.^38,39^

Survey-weighted logistic regression was used to estimate the age-race-sex adjusted odds ratio of poor health status for each disease combination, account for clustering and stratification. Adjustment variables were age as a continuous variable with splines, race (Hispanic, non-Hispanic white, non-Hispanic black, non-Hispanic Asian, and other), and sex. Subgroup analysis was performed by sex as a secondary analysis.

Data pre-processing was performed using SAS version 9.4 for Windows. The analysis was conducted using R version 4.1.0, RStudio (v. 1.4.1717), and R packages: arules (v. 1.6-8) and survey (v. 4.1-1).^40,41^

## RESULTS

The total weighted study population was 249.2 million adults age 18 and older, with 82.0 million (32.9%) reporting receiving treatment for two or more conditions (Table 1) in a single year. 51.7 million (20.7%) adults have three or more conditions, and 30.6 million (12.3%) have 4 or more conditions. A dose response increase is evident between number of chronic conditions and the percentage reporting poor perceived health and average annual medical expenditures. The prevalence of multimorbidity is highest in those ages 65 and older (73.0% or 38.0 million people), followed by age 40 to 64 (35% or 36.4 million people), and 18-39 (7.9% or 7.5 million).

**Table 1:** Number of Chronic Conditions Treated among U.S. Adults by Population Characteristics. Survey-weighted estimates from pooling the 2016-2019 Medical Expenditure Panel Survey. Weighted estimates have been annualized by taking the average over the four year study period. Row percentages are reported under each count of conditions column. Medical expenditures are reported in 2019 US Dollars (USD).

| Variable | No. in unweighted sample | Weighted No. of Adults | 0 chronic conditions,<br>Row % | 1+ chronic conditions,<br>Row % | 2+ chronic conditions,<br>Row % | 3+ chronic conditions,<br>Row % | 4+ chronic conditions,<br>Row % |
| --- | --- | --- | --- | --- | --- | --- | --- |
| Total | 89,947 | 249,221,000 | 47.4% | 52.6% | 32.9% | 20.7% | 12.3% |
| Age Group |  |  |  |  |  |  |  |
| 18 - 39 | 17,745 | 94,573,000 | 73.9% | 26.1% | 7.9% | 2.7% | 0.9% |
| 40 - 64 | 19,776 | 102,614,000 | 41.1% | 58.9% | 35.5% | 20.0% | 10.6% |
| 65 and older | 9,311 | 52,034,000 | 11.6% | 88.4% | 73.0% | 54.9% | 36.1% |
| Sex |  |  |  |  |  |  |  |
| Male | 21,671 | 120,330,000 | 52.4% | 47.6% | 29.9% | 18.8% | 10.7% |
| Female | 25,161 | 128,891,000 | 42.7% | 57.3% | 35.7% | 22.5% | 13.7% |
| Race / Ethnicity |  |  |  |  |  |  |  |
| Hispanic | 12,841 | 40,533,000 | 63.3% | 36.7% | 19.5% | 11.6% | 6.2% |
| Non-Hispanic White only | 21,164 | 156,551,000 | 40.7% | 59.3% | 38.4% | 24.5% | 14.9% |
| Non-Hispanic Black only | 8,113 | 29,523,000 | 53.0% | 47.0% | 28.9% | 18.0% | 10.1% |
| Non-Hispanic Asian only | 3,355 | 15,133,000 | 62.4% | 37.6% | 20.0% | 11.3% | 5.4% |
| Other or Multiple Race | 1,359 | 7,481,000 | 47.8% | 52.2% | 32.7% | 21.0% | 12.8% |
| Self-rated Poor Health | 7,201 | 31,085,000 | 19.0% | 81.0% | 64.6% | 48.8% | 34.3% |
| Per capita medical expenditures in 2019 USD (Median, IQR) | --- | --- | \$322 (\$0 – \$1,382) | \$4,176 (\$1,523 – \$11,067) | \$6,208 (\$2,558 – \$14,858) | \$8,177 (\$3,610 – \$18,756) | \$10,821 (\$5,045 – \$23,712) |
| Per capita out-of-pocket medical expenditures in 2019 USD (Median, IQR) | | | \$30 (\$0 – \$287) | \$542 (\$167 – \$1,401) | \$695 (\$243 – \$1,681) | \$808 (\$292 – \$1,865) | \$903 (\$333 – \$2,080) |

Frequent pattern mining yielded 223 unique combinations of chronic disease, including 74 two-way, 115 three-way, and 34 four-way combinations that affect one million or more of the weighted study population. There are eight combinations that affect 10 million or more adults, 32 combinations that affect 5 million or more adults, and 129 that affect 2 million or more adults per year. There were not any five-way or higher combinations that met the minimum prevalence threshold. The full table is included in the supplementary material (Supplementary Table 2).

Figure 1 shows the most prevalent combinations along with their associated total medical expenditures (in billions of U.S. Dollars) and percent of the population reporting poor perceived health. An estimated 30.8 million people (12.4%, 95%CI: 12.0 - 12.8%) of the U.S. adult population are treated for hypertension and hyperlipidemia – the most common combination (Table 2). An estimated 11.5 million people (4.6%, 95%CI: 4.4% – 4.9%) have hypertension, hyperlipidemia, and diabetes – the most common 3-way combination, and fifth most common combination overall. Hypertension, hyperlipidemia, musculoskeletal disorder, diabetes and arthritis was the most common 5-way combination (1.1%, 95%CI: 1.0% – 1.2%). Combinations including hypertension and/or hyperlipidemia plus another condition comprise the majority of the most prevalent combinations. An alternate version of the analysis excluding hypertension and hyperlipidemia is included in the supplement (Supplementary Figure 2).

**Figure 1:**
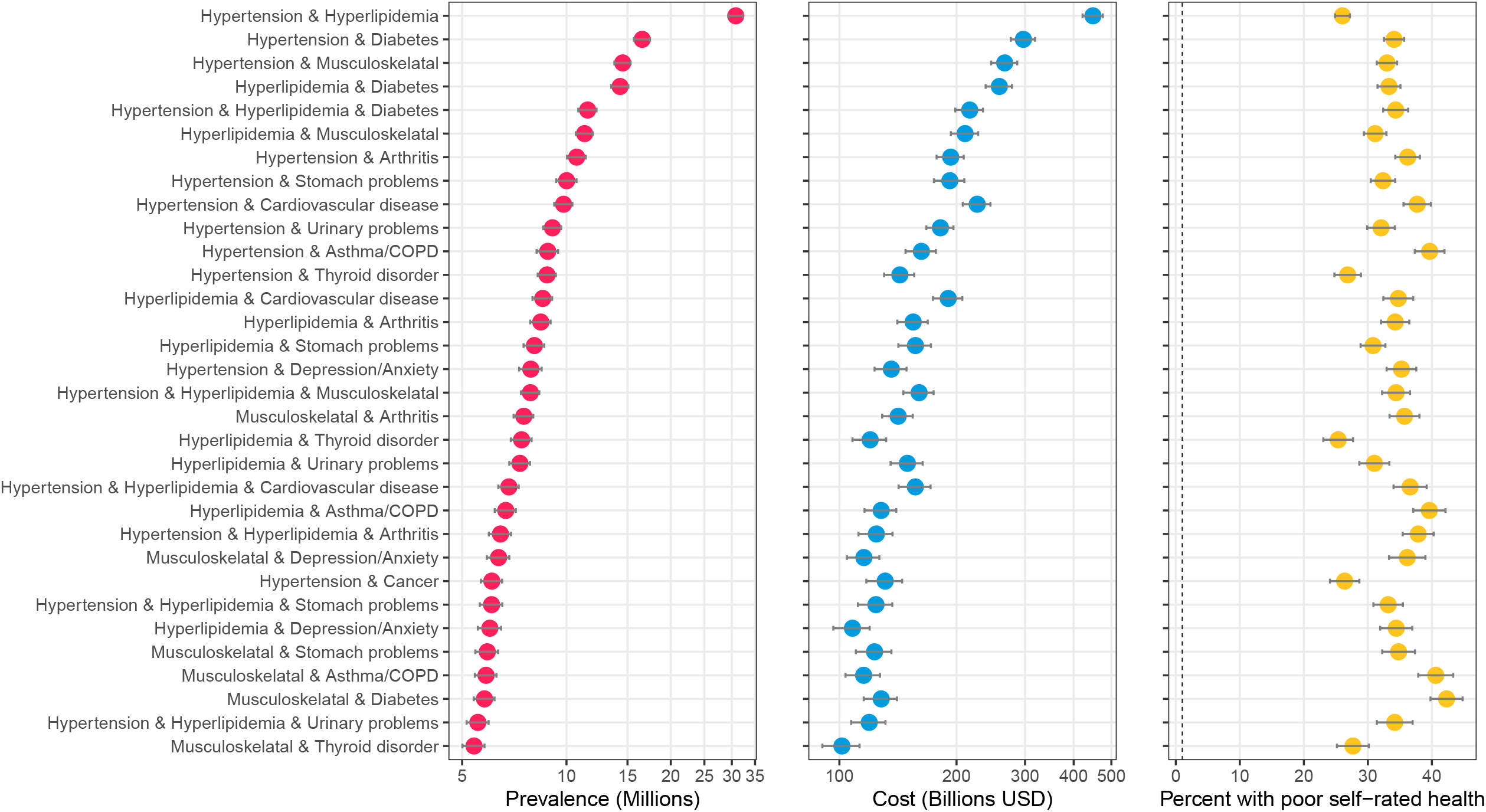
Highly prevalent specific combinations of multimorbidity and associated cost and perceived poor health. The figure shows combinations of conditions with an estimated prevalence of 5 million or more adults in the United States. Data source: the 2016-2019 Medical Expenditure Panel Survey (MEPS). Weighted estimates and 95% confidence intervals were calculated by applying population weights using Taylor-series methods (i.e. complex survey methods) to account for the complex sampling design of the MEPS. Weighted estimates have been annualized by taking the average over the four-year study period. X-axes for prevalence and cost are on the logarithmic scale. Results are sorted by prevalence. Cost is measured as total annual medical expenditures in 2019 US Dollars, and perceived health is an answer of “fair” or “poor” on a 5-point question asking respondents to rank their overall health.

All 223 combinations identified as highly prevalent had O/E ratios (and 95% confidence intervals) greater than 1, indicating the coexistence of these two conditions is unlikely due to chance alone. The combination of hypertension, hyperlipidemia, diabetes, and cardiovascular disease had the highest observed-to-expected ratio of 38.6 (36.4 - 40.9). Cardiovascular disease and heart failure had the highest O/E of 7.4 (95% CI: 6.8 – 8.2%) for dyad combinations and hypertension, cardiovascular disease, and diabetes had the highest O/E of 11.4 (10.8 - 12.0) for triad conditions.

Figure 2 shows specific combinations of multimorbidity that ranked in either the top 20 in terms of average medical expenditures or perceived poor health. A total of 28 combinations met either criteria, and 12 met both. The combination of heart failure and cardiovascular disease had the highest average annual medical expenditures per capita ($33,451, 95%CI: $29,034 – $ 37,868). This was 2.4 times higher than the average for having any two or more chronic conditions ($13,907). Average expenditures were higher than median expenditures across the board, indicating the right-skewed nature of expenditures. The combination of diabetes, asthma/COPD, and arthritis was most associated with perceived poor health status with an adjusted odds ratio of 6.9 (5.4 – 8.8) (Figure 2). The 62.9% reporting poor health was 2.1 times higher than the average percent reporting poor health among those with any three or more chronic conditions. Supplementary Table 2 provide additional details on the out-of-pocket expenses associated with each combination, and Supplemental Figure 3 shows combinations with the highest average out-of-pocket expenditures.

**Figure 2:**
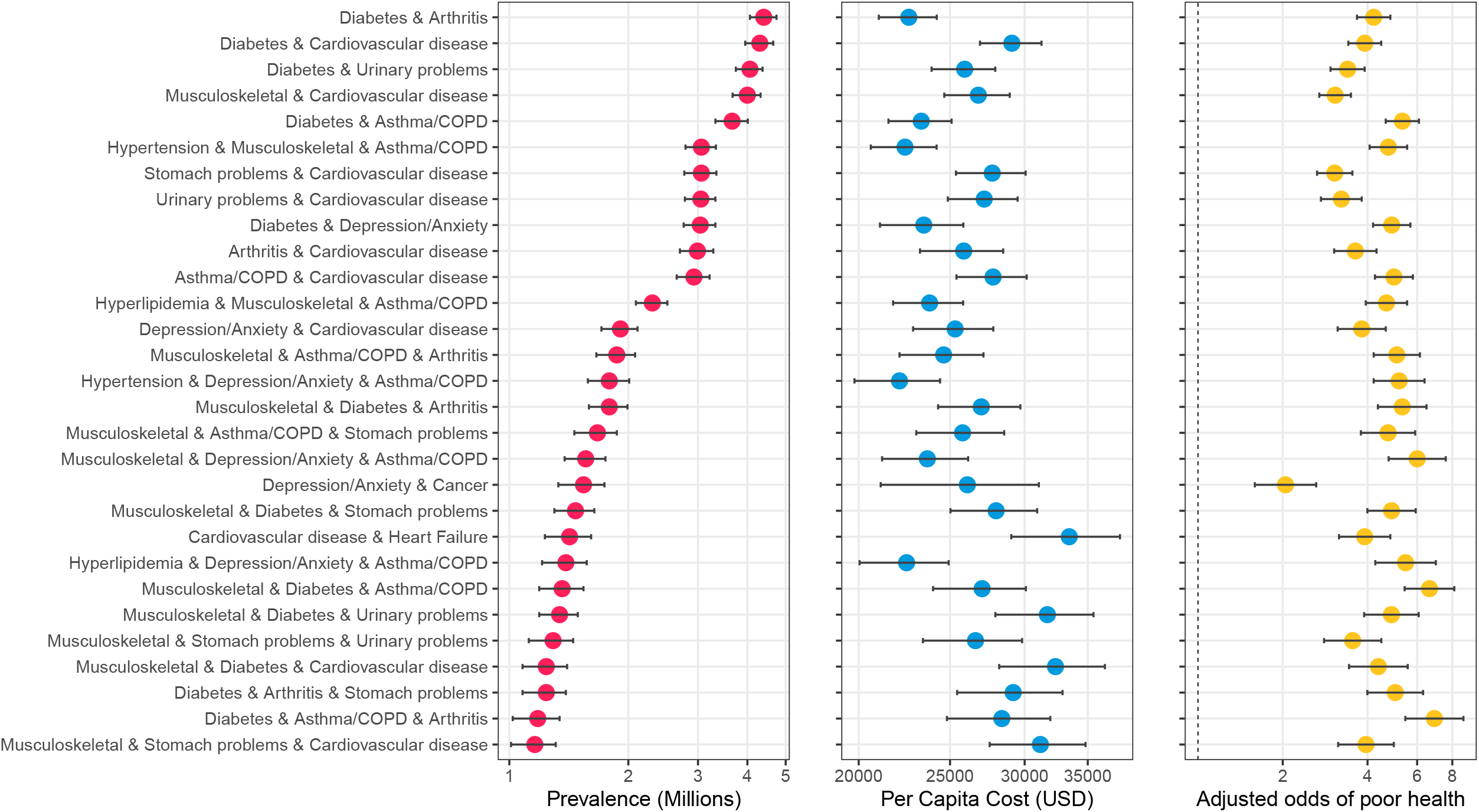
Specific combinations of multimorbidity ranked highest in terms of average cost or perceived poor health. The figure shows the specific combinations of multimorbidity that ranked in the top 20 in terms of highest average cost per person or top 20 highest percentage with self-reported poor health. A total of 28 conditions met either criterion, with 12 meeting both. Data source: 2016-2019 Medical Expenditure Panel Survey (MEPS). Weighted estimates and 95% confidence intervals were calculated by applying population weights using Taylor-series methods (i.e. complex survey methods) to account for the complex sampling design of the MEPS. Weighted estimates have been annualized by taking the average over the four-year study period. Adjusted odds ratios were calculated using log-binomial models adjusting for age, race, and sex, and Taylor-series methods for standard errors. X-axes are on the logarithmic scale, and results are sorted by prevalence.

The top ten combinations among male and female adults in the study population is shown in Figure 3. Hypertension and hyperlipidemia was the top combination for both, but after that the results diverge. Hypertension and diabetes was 2^nd^ for males, but 4^th^ for females. Hypertension and chronic musculoskeletal conditions were 2^nd^ for females, but 4^th^ for males. Combinations with urinary problems appeared twice in the top ten ranking for males, but not for females. Combinations with arthritis, thyroid disorders, stomach problems, and asthma/COPD appeared in the top ten for females but not males.

**Figure 3:**
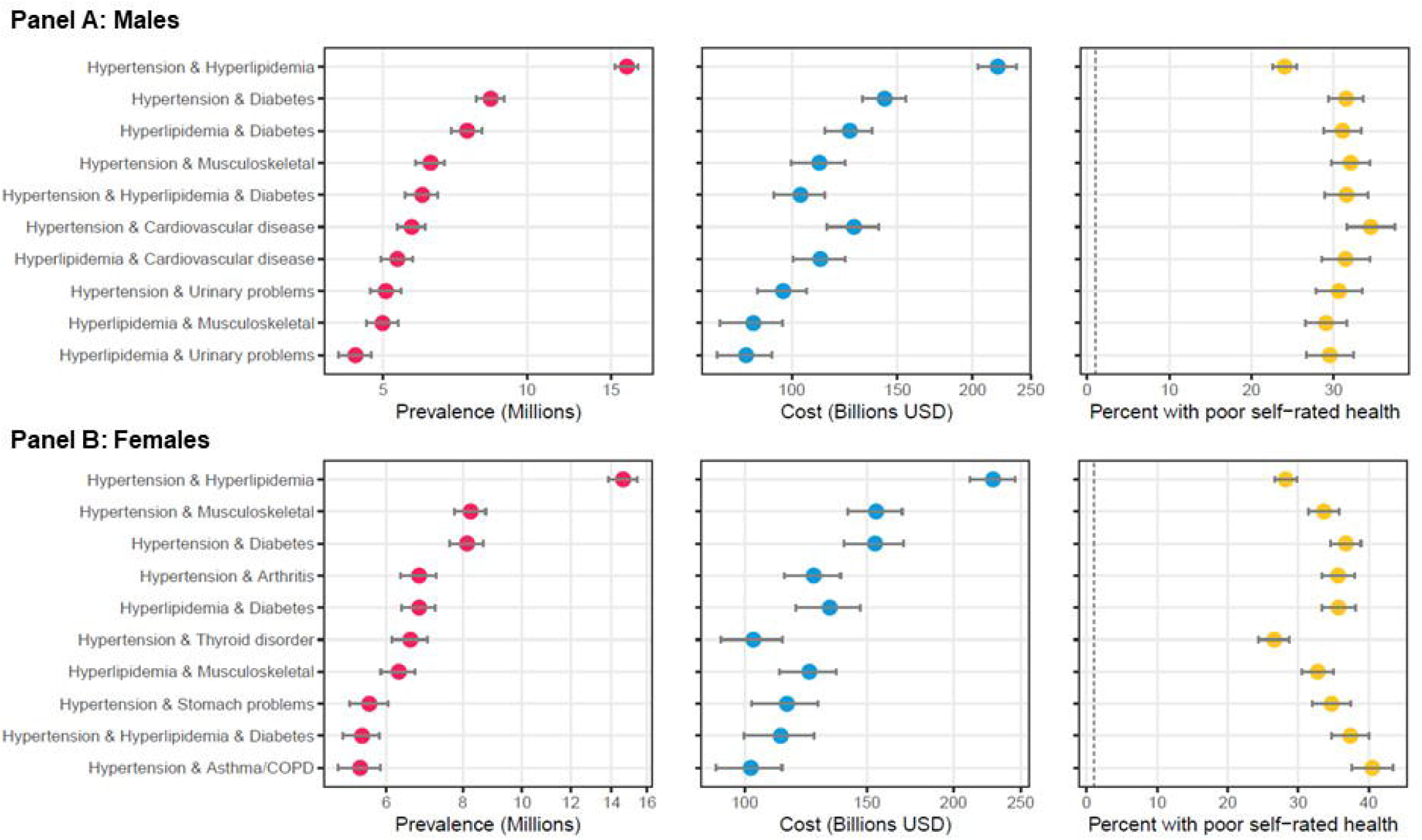
Top ten most prevalent specific combinations of multimorbidity by sex and associated cost and perceived poor health. The figure shows the top ten combinations of conditions as ranked by prevalence by sex among adults in the United States. Panel A shows data on males, and Panel B shows data on females. Data source: the 2016-2019 Medical Expenditure Panel Survey (MEPS). Weighted estimates and 95% confidence intervals were calculated by applying population weights using Taylor-series methods (i.e. complex survey methods) to account for the complex sampling design of the MEPS. Weighted estimates have been annualized by taking the average over the four-year study period. X-axes for prevalence and cost are on the logarithmic scale. Results are sorted by prevalence. Cost is measured as total annual medical expenditures in 2019 US Dollars, and perceived health is an answer of “fair” or “poor” on a 5-point question asking respondents to rank their overall health.

## DISCUSSION

This study combined data mining with survey epidemiology to comprehensively identify and estimate the treatment prevalence of the most frequent combinations of multimorbidity in the United States. This analysis showed that certain multimorbidity combinations of disease are not idiosyncratic, but actually quite common in the US, occurring in one million or more adults.

This paper adds several new contributions to the literature. First, we provide important public health data on the prevalence and associated outcomes of specific multimorbidity combinations. While it was not surprising to find hypertension and hyperlipidemia as the top combination, a fact that is well-established,^42^ the approximate ranking and estimated prevalence of many other combinations farther down the list was previously unknown. Second, this study provides data on observed-to-expected prevalence ratio for each combination. Disease with high ratios likely share risk factors or have common pathways for disease, which could spur hypotheses for future research. Finally, we identify combinations with the highest burden in terms of total expenditures and perceived poor health. These combinations could be priority targets for future prevention or treatment interventions targeted toward multimorbidity patients.

Despite the high prevalence of these disease combinations, most evidence-based practice guidelines are single-disease focused.^43^ This relegates managing people with multimorbidity, who represent the majority of people presenting for primary care,^44,45^ to the “art”, rather than the science of medicine, and has led to largely untargeted global interventions that show limited or no effect in clinical trials.^46^ While general approaches for complex patients exist including geriatric care teams,^47^ the Age-Friendly Health Systems 4M model of care,^48^ and deprescribing guidelines (e.g. Beers Criteria and STOPP/START),^49,50^ significant evidence gaps remain especially when it comes to specific guidelines for managing patients with specific combinations of conditions. For some conditions, multimorbidity may be the norm. For example, this study showed that 45 million adults are treated for hyperlipidemia, and of those approximately two-thirds are treated for hypertension as well. This means it is more common for a patient to be treated for high cholesterol plus another condition, than just managing cholesterol alone.

The highly prevalent combinations identified in this study could be prioritized for future evidence-based research. Some combinations show the possibility that a single medication could target multiple diseases thus reducing the polypharmacy. For example, a low-dose antidepressant for anxiety, can also treat pain from arthritis or diabetic neuropathy. Other common combinations show the potential of physical activity, diet, and tobacco cessation interventions to positively affect multiple conditions and reduce medication burden.

This was, to the author’s knowledge, the most comprehensive, population-based study in the U.S. on the prevalence and associated impact of multimorbidity combinations in terms of the number of conditions, the examination of higher-order combinations (e.g. 4-way or deeper), and the broadness of the study population. Ward and Schiller examined common dyad and triad combinations of ten conditions using the National Health Interview Survey.^23^ The most common dyad condition was hypertension-arthritis and 26% of adults had multimorbidity. A similar more recent study using 2018 NHIS data estimated that 27.2% of U.S. adults had two or more.^51^ Weiss et al, used survey weights and examined deeper combinations of chronic disease, but only in five conditions using 1999-2004 National Health and Nutrition Examination Survey (NHANES) data.^24^

Several studies have been conducted in specific subpopulations of the U.S. using ARM or similar methods to examine combinations of multimorbidity. Koroukian et al. used ARM to identify combinations of multimorbidity associated with high expenditures among Medicare enrollees.^26^ Ho et al. used ARM to identify high-risk comorbidity combinations for patients undergoing emergency general surgery using the Nationwide Inpatient Sample.^38^ Quinones et al. (2016) and Quinones et al. (2019) examined diabetes comorbidities and racial disparities in multimorbidity, respectively using the Health & Retirement Study.^17,52^ Quinones et al. (2022) examined combinations associated of multimorbidity among patients seeking care at community health centers.^53^ Steinman et al. examined combinations of multimorbidity impacting older veterans using data form the Veterans Affairs health system.^21^

Comparable studies have been conducted internationally outside the U.S., including a study using the Korea Health Panel which found an overall multimorbidity rate of 34.8% in South Korea,^54^ and a study using Beijing medical claims data in China, which found a higher rate of multimorbidity than our study (51.6% in mid-life adults and 81.3% in older adults). This is probably due to including more conditions. Zemekidun et al. applied cluster analysis and ARM to the UK biobank to identify combinations of multimorbidity.^55^ Britt et al. examined prevalence and patterns of multimorbidity in patient-reported surveys in Australia across nine morbidity domains.^56^ The most common combination was arthritis/chronic back pain + vascular disease (15% of population). Nicholson et al. developed a cluster analysis tool to identify common combinations and sequences in Canada using an approach that seems similar to association rule mining.^32^

### Strengths

The data mining allowed a more comprehensive identification of high frequency multimorbidity combinations than previous efforts.^23,24^ This study uses a national sample, representative of the U.S. population. The MEPS database is also unique among national surveys in that its open-ended condition enumeration approach (rather than a fixed set of questions) allows for a more comprehensive capture of different chronic conditions, compared to other national surveys.^17,23,24^ MEPS covers all payers (even uninsured) and has data on services from all providers, which is an advantage over big data sources like claims data and electronic health records, respectively.

Association rule mining allows for the identification of highly prevalent combinations in a quick and computationally efficient manner, while filtering out combinations that have no observations or are sparse. While our study only focused on combinations occurring in over 1 million adults, the method could also be used to identify higher order combinations by setting a lower prevalence thresholds. Association rule mining is still relatively uncommon in clinical and epidemiological studies. The method could be applied to other areas of research, for example identifying high-risk combinations of risk factors associated with mortality,^38^ social determinant of health indicators associated with disparities,^57^ or prescription drug combinations associated with adverse drug events.^58^ To our knowledge, this is the first study to combine complex survey analysis with association rule mining to yield national estimates of combinations of variables.

### Limitations

MEPS provides accurate estimates of treated prevalence for chronic disease, but this is likely lower than the underlying population prevalence for some conditions, especially those that are less salient.^30^ On the other hand, the focus on treated conditions represent those for which care decisions between multiple conditions may conflict. The MEPS includes community-dwelling subjects only, meaning institutionalized persons such as nursing home and assisted living residents are excluded. Estimates of total expenditures are typically lower than what is estimated from the National Health Expenditure Accounts (NHEA), because the MEPS does not include these institutionalized populations.^59^ ICD-10 codes were classified into 20 primary-care relevant conditions using an established algorithm, but other classification systems may result in different combinations and prevalence estimates.^31^ This algorithm was chosen over others because it was originally designed for use on self-reported conditions, and that it could be applied with the 3-digit ICD-10 codes available in MEPS. Many other algorithms require the use of fully-specified ICD-10 codes and/or were designed for use in administrative data, including Elixhauser comorbidities,^60^ Deyo-Charlson Comorbidity Index,^61,62^ Clinical Classification Software,^63^ and Phecodes.^64^ Consensus on which conditions to include in measures of multimorbidity has not been established, and is a problem that continues to plague the field.^65^

This study used the most recent data (2016-2019) available at the time; therefore, the impact of the COVID-19 pandemic was not captured in these results. As comorbidities were among the greatest risk factors of COVID-related death, excess mortality in people with multimorbidity could have lowered population prevalence rates.^66–68^ However, there is growing evidence of increases in prevalence of certain post-Covid conditions (i.e. long COVID) including respiratory problems, diabetes, heart conditions, neurological conditions, migraines, and mental health conditions.^69–73^ Understanding the scale and impact of post-COVID conditions is an on-going area of investigation in the scientific community.

## Conclusion

This cross-sectional analysis of a nationally-representative survey showed that certain multimorbidity combinations of disease are quite common in the US, occurring in one million or more adults. The combinations reported here could be prioritized for evidence-based research and integration into practice guidelines, especially those most associated with poor health and high medical costs.

## Data Availability

This study uses openly available data from the Medical Expenditure Panel Survey, sponsored by the US Department of Health & Human Services, Agency for Healthcare Research & Quality

https://www.meps.ahrq.gov/mepsweb/data_stats/download_data_files.jsp

## DATA AVAILABILITY STATEMENT

This study uses openly available data from the Medical Expenditure Panel Survey. Available at: https://www.meps.ahrq.gov/mepsweb/data_stats/download_data_files.jsp. Requests to access the processed dataset used in analysis should contact the corresponding author. Project code is posted on the author’s GitHub: https://github.com/nickschiltz/multimorbidity-prevalence

## ETHICS STATEMENT

This study uses only publicly available deidentified datasets. The Case Western Reserve University Institutional Review Board reviewed the protocol for this study and deemed it to be exempt under U.S. federal law.

## AUTHOR CONTRIBUTIONS

NKS is the sole author, had full access to the data, and takes responsibility for the integrity of the data and the accuracy of the data analysis, and the entirety of the manuscript. NKS contributions include conceptualization, design of the study, statistical analysis, interpreting the data, and writing and revising the manuscript.

## FUNDING

This project was supported in part by NIH/NCATS CTSA KL2TR002547. Its contents are solely the responsibility of the author and do not necessarily represent the official views of the NIH.

## CONFLICTS OF INTEREST

The author of this report has no conflicts of interest to disclose.

## ACKNOWLEDGEMENTS

The author would like to thank Obada Farhan, MS (Case Western Reserve University) and R. Henry Olaisen for their roles in writing some of the R code used in the analysis, and Kurt Stange, MD, PhD for his mentorship on the career development award that supported this work.

## Notes

### Competing Interest Statement

The authors have declared no competing interest.

### Funding Statement

This project was supported in part by NIH/NCATS CTSA KL2TR0002547.

### Author Declarations

This study uses openly available data from the Medical Expenditure Panel Survey. Available at: https://www.meps.ahrq.gov/mepsweb/data_stats/download_data_files.jsp

### Summary of Updates

Table 2 removed (redundant with supplemental table 2) Figure 3 added (sex differences) Introduction expanded Discussion expanded Edits to methods and results Supplementary materials updated

