## Supplemental Materials for "Prevalence of multimorbidity combinations and their association with medical costs and poor health: a population-based study of U.S. adults"

**Supplemental Figure 1: Flow Diagram of Study Population**

**Figure 1 caption:** Flow diagram of how the sample was selected from the original Medical Expenditure Panel Study data. Because MEPS involves two-year overlapping cohorts, some subjects appear in the database twice. We follow the methods recommended by MEPS for pooling multiple years of data, including complex survey design analysis controlling for clustering and stratification. *exact count of subjects with missing data is masked to comply with data use agreement requiring cell counts less than 10 to be masked.

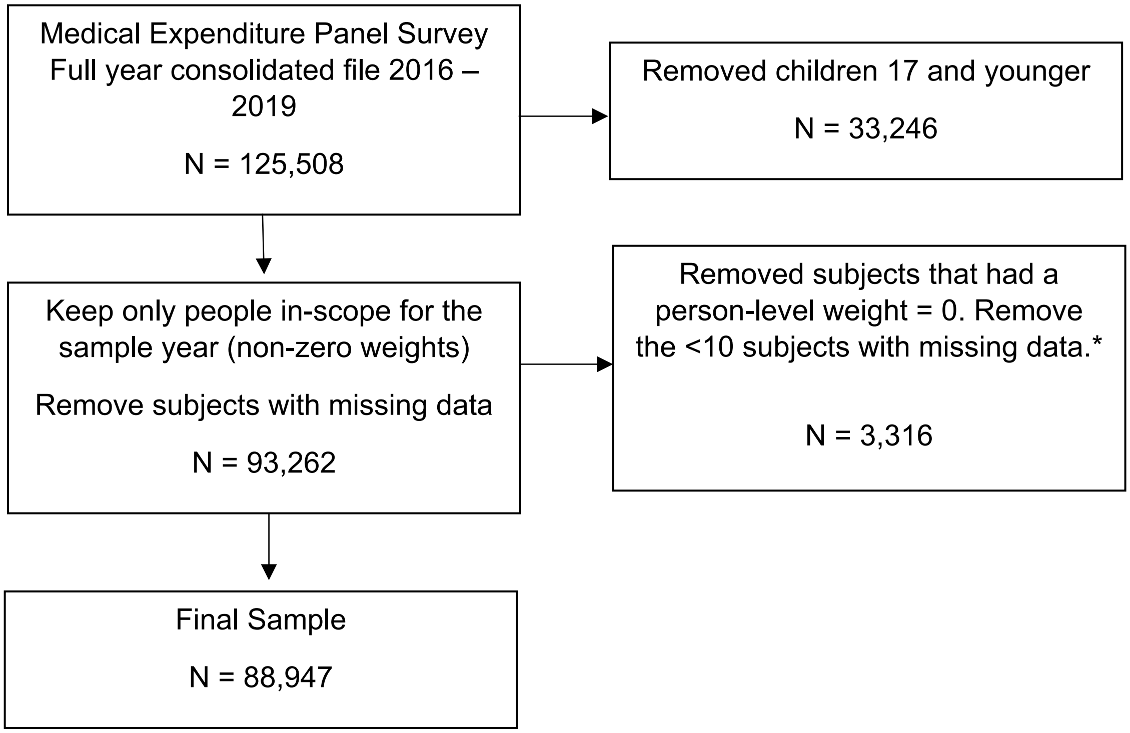

### Supplementary Table 1: Algorithm used to classify primary care relevant conditions

| ICD-10-CM Codes | Label | Notes |
|---|---|---|
| I10–I15 | Hypertension (high blood pressure) | |
| F33, F40, F41 | Depression or anxiety | |
| M40–M54, M60–M63, M65–M68, M70–M79 | Chronic musculoskeletal conditions causing pain or limitation | |
| M05, M13, M15–M19 | Arthritis and/or rheumatoid arthritis | *original paper used only subcodes M05.9, M13.0, and M13.9 of M05 and M13, respectively. |
| M81 | Osteoporosis | |
| J40–J46 | Asthma, chronic obstructive pulmonary disease (COPD), or chronic bronchitis | |
| I20, I25, I48, I70–I79 | Cardiovascular disease (angina, myocardial infarction, atrial fibrillation, poor circulation in the lower limbs) | |
| I05–I09, I34–I39, I42, I43, I50 | Heart failure (including valve problems or replacement) | |
| G45, I62 | Stroke and transient ischemic attack | |
| K21, K25, K29 | Stomach problem (reflux, heartburn, or gastric ulcer) | *original paper used only subcodes K25.7, K29.5 of K25 and K29, respectively |
| K50–K52, K57, K58 | Colon problem (irritable bowel, Crohn's disease, ulcerative colitis, diverticulosis) | |
| K70–K77 | Chronic hepatitis | |
| E10–E14 | Diabetes | |
| E00–E07 | Thyroid disorder | |
| C00–C97 | Cancer | |
| N18,N19 | Kidney disease or failure | |
| N03, N11, N18, N20–N23, N25–N29, N30–N39, N40–N51 | Chronic urinary problem | |
| F00-F03 | Dementia or Alzheimer's disease | |
| E78 | Hyperlipidemia (high cholesterol) | |
| E66 | Obesity | |

**Supplementary Table 1 Caption:** Each medical condition reported in the MEPS is captured from an open-ended question and the verbatim responses are coded to ICD-10-CM code. The ICD-10-CM were mapped to one of 20 primary care relevant chronic conditions as shown in the table based on an algorithm published by Fortin et al.[31] Some modifications were required from the published algorithm as noted, due to the MEPS only including 3-digit ICD-10-CM codes in public release files.

### Supplementary Table 2: Specific Multimorbidity Combinations among U.S. Adults, 2016 – 2019

**Supplementary Table 2 Caption:** Specific multi-way combinations of chronic disease ranked by prevalence from the 2016-2019 Medical Expenditure Panel Survey (MEPS). Weighted number and percent and 95% confidence intervals were calculated by applying population weights using Taylor-series methods (i.e. complex survey methods) to account for the complex sampling design of the MEPS. Weighted estimates have been annualized by taking the average over the four-year study period. The observed to expected ratio (also known as lift) is the observed prevalence of the combination divided by the expected prevalence of the combination if each disease within the combination were statistically independent. Total expenditures and total out-of-pocket expenditures are reported in billions of US dollars and adjusted for inflation (reported in 2019 USD). Percent reporting poor health are those that responded "fair" or "poor" on a five point question asking respondents to rank their overall health.

| Overall Rank (CI) | Multimorbidity Combinations | Prevalence, % (95% CI) | Weighted No. of Adults in millions (95% CI) | Total Expenditures in billions USD (95% CI) | Total out-of-pocket burden in billions USD (95% CI) | Percent Reporting Poor Health (95% CI) | Observed to Expected Ratio (95% CI) |
|---|---|---|---|---|---|---|---|
| **Single conditions** | | | | | | | |
| 1 (1 - 1) | Hypertension | 24.46 (23.87 - 25.06) | 60.96 (58.33 - 63.60) | 773 (734 - 813) | 85.1 (79.5 - 90.6) | 23.5 (22.7 - 24.3) | 1.0 |
| 2 (2 - 2) | Hyperlipidemia | 18.08 (17.59 - 18.57) | 45.05 (43.00 - 47.09) | 602 (568 - 636) | 65.0 (60.5 - 69.5) | 22.9 (22.0 - 23.8) | 1.0 |
| 3 (3 - 3) | Musculoskeletal | 14.86 (14.45 - 15.29) | 37.04 (35.28 - 38.81) | 503 (472 - 533) | 59.7 (55.2 - 64.2) | 23.7 (22.7 - 24.7) | 1.0 |
| 5 (5 - 5) | Diabetes | 9.81 (9.46 - 10.17) | 24.45 (23.26 - 25.64) | 399 (373 - 425) | 35.8 (33.0 - 38.5) | 32.0 (30.7 - 33.4) | 1.0 |
| 6 (6 - 6) | Depression/Anxiety | 8.95 (8.62 - 9.29) | 22.31 (21.12 - 23.49) | 294 (273 - 315) | 34.5 (31.3 - 37.6) | 25.6 (24.1 - 27.0) | 1.0 |
| 7 (7 - 8) | Asthma/COPD | 8.27 (7.97 - 8.57) | 20.60 (19.44 - 21.76) | 289 (267 - 311) | 29.6 (27.2 - 32.0) | 29.7 (28.1 - 31.4) | 1.0 |
| 8 (7 - 8) | Thyroid disorder | 8.17 (7.85 - 8.49) | 20.36 (19.20 - 21.51) | 257 (238 - 277) | 35.2 (31.5 - 39.0) | 20.2 (18.9 - 21.5) | 1.0 |
| 9 (9 - 10) | Arthritis | 7.40 (7.09 - 7.71) | 18.44 (17.47 - 19.41) | 300 (278 - 321) | 30.7 (28.2 - 33.3) | 30.5 (29.1 - 32.0) | 1.0 |
| 10 (9 - 11) | Urinary problems | 7.30 (7.03 - 7.58) | 18.20 (17.23 - 19.17) | 296 (275 - 316) | 32.2 (29.2 - 35.3) | 24.4 (23.1 - 25.8) | 1.0 |
| 11 (9 - 11) | Stomach problems | 7.06 (6.74 - 7.40) | 17.60 (16.57 - 18.64) | 295 (272 - 319) | 29.2 (26.3 - 32.1) | 27.7 (26.3 - 29.2) | 1.0 |
| 15 (13 - 15) | Cardiovascular disease | 5.65 (5.42 - 5.90) | 14.09 (13.30 - 14.88) | 313 (289 - 337) | 27.2 (24.6 - 29.7) | 34.4 (32.6 - 36.2) | 1.0 |
| 16 (16 - 18) | Cancer | 4.69 (4.45 - 4.94) | 11.69 (10.96 - 12.41) | 233 (212 - 254) | 22.4 (20.5 - 24.4) | 22.5 (20.9 - 24.2) | 1.0 |
| 61 (57 - 70) | Colon problems | 1.50 (1.38 - 1.62) | 3.73 (3.42 - 4.05) | 65 (58 - 73) | 7.2 (6.1 - 8.2) | 28.0 (24.8 - 31.3) | 1.0 |
| 77 (71 - 90) | Stroke | 1.29 (1.18 - 1.40) | 3.21 (2.91 - 3.51) | 76 (66 - 86) | 5.4 (4.5 - 6.3) | 42.6 (39.2 - 46.0) | 1.0 |
| 79 (71 - 90) | Heart Failure | 1.28 (1.18 - 1.39) | 3.19 (2.91 - 3.47) | 82 (72 - 93) | 6.4 (5.4 - 7.4) | 38.5 (34.7 - 42.4) | 1.0 |
| 150 (134 - 168) | Osteoporosis | 0.77 (0.69 - 0.86) | 1.92 (1.70 - 2.14) | 32 (26 - 38) | 4.4 (2.8 - 6.0) | 20.1 (16.3 - 23.9) | 1.0 |
| 235 (212 - 240) | Hepatitis | 0.48 (0.42 - 0.54) | 1.19 (1.03 - 1.35) | 25 (18 - 32) | 2.0 (1.6 - 2.5) | 41.8 (36.0 - 47.7) | 1.0 |
| n/a | Obesity | 0.46 (0.40 - 0.53) | 1.15 (0.98 - 1.33) | 21 (16 - 26) | 2.0 (1.3 - 2.7) | 26.9 (21.5 - 32.2) | --- |
| n/a | Kidney disease | 0.46 (0.40 - 0.52) | 1.14 (0.98 - 1.30) | 50 (40 - 60) | 2.4 (1.9 - 3.0) | 50.1 (43.7 - 56.5) | --- |
| n/a | Dementia or Alzheimer's | 0.35 (0.30 - 0.41) | 0.87 (0.73 - 1.00) | 19 (15 - 23) | 2.8 (1.4 - 4.2) | 46.8 (39.6 - 54.0) | --- |

| | | | | | | | |
|---|---|---|---|---|---|---|---|
| **Dyad (2-way) multimorbidity combinations** | | | | | | | |
| 4 (4 - 4) | Hypertension & Hyperlipidemia | 12.36 (11.96 - 12.77) | 30.80 (29.33 - 32.28) | 448 (422 - 475) | 45.8 (42.2 - 49.4) | 26.0 (24.9 - 27.2) | 2.7 (2.6 - 2.7) |
| 12 (12 - 12) | Hypertension & Diabetes | 6.64 (6.35 - 6.93) | 16.54 (15.65 - 17.42) | 297 (276 - 318) | 25.5 (23.2 - 27.8) | 34.1 (32.5 - 35.7) | 2.6 (2.5 - 2.7) |
| 13 (13 - 15) | Hypertension & Musculoskeletal | 5.83 (5.58 - 6.08) | 14.53 (13.75 - 15.31) | 266 (245 - 286) | 26.6 (23.8 - 29.4) | 33.0 (31.4 - 34.6) | 1.6 (1.6 - 1.7) |
| 14 (13 - 15) | Hyperlipidemia & Diabetes | 5.73 (5.47 - 6.00) | 14.28 (13.47 - 15.09) | 258 (238 - 278) | 22.5 (20.5 - 24.5) | 33.3 (31.5 - 35.1) | 3.1 (3.0 - 3.2) |
| 18 (16 - 18) | Hyperlipidemia & Musculoskeletal | 4.53 (4.33 - 4.73) | 11.28 (10.64 - 11.91) | 210 (193 - 227) | 21.2 (18.7 - 23.6) | 31.1 (29.4 - 32.9) | 1.7 (1.6 - 1.8) |
| 19 (18 - 19) | Hypertension & Arthritis | 4.29 (4.07 - 4.53) | 10.70 (10.04 - 11.37) | 193 (178 - 209) | 17.9 (16.2 - 19.6) | 36.2 (34.3 - 38.1) | 2.3 (2.3 - 2.4) |
| 20 (20 - 21) | Hypertension & Stomach problems | 4.02 (3.79 - 4.26) | 10.01 (9.34 - 10.69) | 192 (175 - 209) | 18.0 (15.7 - 20.3) | 32.4 (30.4 - 34.3) | 2.3 (2.2 - 2.4) |
| 21 (20 - 21) | Hypertension & Cardiovascular disease | 3.94 (3.74 - 4.14) | 9.81 (9.22 - 10.40) | 226 (208 - 244) | 18.7 (16.9 - 20.6) | 37.7 (35.6 - 39.8) | 2.7 (2.6 - 2.8) |
| 22 (22 - 24) | Hypertension & Urinary problems | 3.66 (3.48 - 3.84) | 9.11 (8.57 - 9.65) | 182 (167 - 196) | 17.1 (15.5 - 18.8) | 32.1 (29.9 - 34.2) | 2.0 (2.0 - 2.1) |
| 23 (22 - 26) | Hypertension & Asthma/COPD | 3.54 (3.34 - 3.75) | 8.83 (8.20 - 9.46) | 162 (148 - 177) | 14.6 (13.1 - 16.1) | 39.6 (37.3 - 42.0) | 1.8 (1.7 - 1.8) |
| 24 (22 - 26) | Hypertension & Thyroid disorder | 3.53 (3.35 - 3.71) | 8.79 (8.26 - 9.32) | 143 (130 - 156) | 18.1 (15.5 - 20.8) | 26.8 (24.8 - 28.9) | 1.8 (1.7 - 1.8) |
| 25 (23 - 27) | Hyperlipidemia & Cardiovascular disease | 3.43 (3.24 - 3.62) | 8.54 (7.98 - 9.09) | 190 (174 - 207) | 15.4 (14.0 - 16.9) | 34.7 (32.4 - 37.1) | 3.2 (3.1 - 3.3) |
| 26 (23 - 27) | Hyperlipidemia & Arthritis | 3.38 (3.18 - 3.59) | 8.43 (7.86 - 9.00) | 155 (141 - 169) | 14.6 (13.0 - 16.2) | 34.3 (32.1 - 36.5) | 2.5 (2.4 - 2.6) |
| 27 (25 - 29) | Hyperlipidemia & Stomach problems | 3.24 (3.05 - 3.44) | 8.08 (7.52 - 8.63) | 157 (142 - 172) | 14.4 (13.0 - 15.8) | 30.8 (28.9 - 32.7) | 2.5 (2.4 - 2.6) |
| 28 (27 - 31) | Hypertension & Depression/Anxiety | 3.17 (2.96 - 3.38) | 7.89 (7.30 - 8.48) | 136 (123 - 149) | 13.2 (11.1 - 15.4) | 35.2 (32.9 - 37.6) | 1.5 (1.4 - 1.6) |
| 30 (28 - 32) | Musculoskeletal & Arthritis | 3.02 (2.85 - 3.20) | 7.53 (7.04 - 8.02) | 142 (129 - 154) | 14.1 (12.6 - 15.6) | 35.7 (33.4 - 38.1) | 2.7 (2.6 - 2.9) |
| 31 (29 - 32) | Hyperlipidemia & Thyroid disorder | 2.98 (2.80 - 3.16) | 7.42 (6.91 - 7.93) | 120 (108 - 132) | 14.5 (12.2 - 16.7) | 25.4 (23.0 - 27.7) | 2.0 (2.0 - 2.1) |
| 32 (30 - 32) | Hyperlipidemia & Urinary problems | 2.95 (2.77 - 3.13) | 7.34 (6.84 - 7.85) | 150 (135 - 164) | 14.3 (12.7 - 15.9) | 31.0 (28.7 - 33.4) | 2.2 (2.1 - 2.3) |
| 34 (33 - 36) | Hyperlipidemia & Asthma/COPD | 2.68 (2.53 - 2.84) | 6.68 (6.21 - 7.14) | 128 (116 - 140) | 11.1 (9.9 - 12.3) | 39.6 (37.1 - 42.1) | 1.8 (1.7 - 1.9) |
| 36 (34 - 39) | Musculoskeletal & Depression/Anxiety | 2.56 (2.39 - 2.73) | 6.37 (5.89 - 6.84) | 116 (105 - 127) | 11.7 (10.3 - 13.2) | 36.1 (33.3 - 39.0) | 2.0 (1.9 - 2.1) |
| 37 (35 - 42) | Hypertension & Cancer | 2.45 (2.30 - 2.60) | 6.09 (5.66 - 6.52) | 131 (117 - 145) | 12.0 (10.7 - 13.4) | 26.4 (24.1 - 28.7) | 2.0 (1.9 - 2.1) |
| 39 (36 - 42) | Hyperlipidemia & Depression/Anxiety | 2.41 (2.26 - 2.58) | 6.01 (5.55 - 6.48) | 108 (96 - 120) | 10.5 (9.2 - 11.8) | 34.4 (31.9 - 36.9) | 1.5 (1.5 - 1.6) |
| 40 (37 - 43) | Musculoskeletal & Stomach problems | 2.37 (2.22 - 2.53) | 5.91 (5.46 - 6.35) | 123 (110 - 136) | 11.8 (10.3 - 13.2) | 34.8 (32.2 - 37.3) | 2.3 (2.2 - 2.4) |
| 41 (37 - 43) | Musculoskeletal & Asthma/COPD | 2.35 (2.21 - 2.50) | 5.86 (5.44 - 6.28) | 115 (104 - 127) | 11.3 (9.9 - 12.7) | 40.6 (37.9 - 43.3) | 2.0 (1.9 - 2.1) |
| 42 (37 - 43) | Musculoskeletal & Diabetes | 2.33 (2.19 - 2.47) | 5.80 (5.40 - 6.20) | 128 (116 - 141) | 10.4 (9.3 - 11.6) | 42.3 (39.8 - 44.8) | 1.6 (1.5 - 1.7) |
| 44 (43 - 44) | Musculoskeletal & Thyroid disorder | 2.17 (2.03 - 2.31) | 5.41 (5.01 - 5.80) | 102 (90 - 113) | 13.2 (11.0 - 15.4) | 27.7 (25.2 - 30.1) | 1.8 (1.7 - 1.9) |
| 45 (45 - 48) | Musculoskeletal & Urinary problems | 2.00 (1.87 - 2.13) | 4.98 (4.62 - 5.35) | 111 (100 - 121) | 11.3 (9.7 - 12.9) | 33.0 (30.2 - 35.9) | 1.9 (1.8 - 2.0) |
| 47 (45 - 48) | Hyperlipidemia & Cancer | 1.95 (1.81 - 2.10) | 4.86 (4.47 - 5.26) | 109 (95 - 123) | 9.7 (8.5 - 10.8) | 24.3 (21.9 - 26.7) | 2.3 (2.1 - 2.4) |
| 49 (49 - 52) | Diabetes & Arthritis | 1.77 (1.64 - 1.90) | 4.40 (4.06 - 4.74) | 99 (89 - 110) | 8.0 (6.9 - 9.2) | 45.4 (42.3 - 48.5) | 2.3 (2.2 - 2.5) |
| 51 (49 - 57) | Diabetes & Cardiovascular disease | 1.73 (1.60 - 1.86) | 4.30 (3.95 - 4.65) | 125 (111 - 139) | 8.7 (7.5 - 9.8) | 44.2 (41.0 - 47.4) | 2.9 (2.8 - 3.1) |

| | | | | | | | |
|---|---|---|---|---|---|---|---|
| 53 (49 - 60) | Arthritis & Stomach problems | 1.64 (1.51 - 1.78) | 4.09 (3.73 - 4.45) | 86 (76 - 96) | 8.1 (6.9 - 9.2) | 37.6 (34.7 - 40.4) | 3.1 (2.9 - 3.3) |
| 54 (49 - 60) | Diabetes & Stomach problems | 1.63 (1.51 - 1.77) | 4.07 (3.73 - 4.42) | 103 (89 - 116) | 7.5 (6.4 - 8.5) | 42.9 (40.0 - 45.8) | 2.2 (2.1 - 2.4) |
| 55 (51 - 60) | Diabetes & Urinary problems | 1.63 (1.51 - 1.75) | 4.06 (3.74 - 4.37) | 105 (93 - 117) | 7.9 (6.9 - 8.9) | 40.8 (37.6 - 44.0) | 2.2 (2.1 - 2.4) |
| 57 (52 - 61) | Musculoskeletal & Cardiovascular disease | 1.60 (1.49 - 1.72) | 4.00 (3.67 - 4.32) | 107 (95 - 119) | 8.5 (7.0 - 10.0) | 39.1 (36.1 - 42.1) | 1.9 (1.8 - 2.0) |
| 58 (52 - 68) | Asthma/COPD & Arthritis | 1.58 (1.45 - 1.71) | 3.93 (3.57 - 4.29) | 81 (71 - 91) | 6.6 (5.7 - 7.5) | 43.6 (40.2 - 46.9) | 2.6 (2.5 - 2.8) |
| 59 (53 - 68) | Depression/Anxiety & Asthma/COPD | 1.56 (1.44 - 1.69) | 3.88 (3.54 - 4.23) | 73 (64 - 82) | 6.4 (5.4 - 7.3) | 42.4 (38.7 - 46.1) | 2.3 (2.1 - 2.4) |
| 60 (53 - 68) | Asthma/COPD & Stomach problems | 1.56 (1.43 - 1.69) | 3.88 (3.52 - 4.24) | 85 (74 - 95) | 7.2 (6.1 - 8.4) | 40.7 (37.4 - 44.1) | 2.8 (2.6 - 2.9) |
| 62 (57 - 72) | Depression/Anxiety & Stomach problems | 1.48 (1.36 - 1.62) | 3.70 (3.35 - 4.04) | 80 (69 - 91) | 7.2 (5.5 - 8.8) | 39.1 (36.0 - 42.2) | 2.4 (2.3 - 2.6) |
| 63 (58 - 72) | Arthritis & Thyroid disorder | 1.48 (1.36 - 1.60) | 3.68 (3.36 - 4.01) | 67 (60 - 75) | 7.8 (6.6 - 8.9) | 31.9 (28.7 - 35.0) | 2.3 (2.2 - 2.5) |
| 64 (57 - 72) | Diabetes & Thyroid disorder | 1.47 (1.34 - 1.61) | 3.67 (3.32 - 4.01) | 76 (65 - 86) | 7.5 (6.3 - 8.7) | 34.7 (31.4 - 38.0) | 1.8 (1.7 - 1.9) |
| 64 (58 - 72) | Diabetes & Asthma/COPD | 1.47 (1.35 - 1.60) | 3.66 (3.32 - 4.01) | 85 (75 - 96) | 6.0 (5.2 - 6.7) | 50.1 (46.8 - 53.5) | 1.8 (1.7 - 1.9) |
| 72 (62 - 82) | Depression/Anxiety & Arthritis | 1.37 (1.26 - 1.49) | 3.42 (3.10 - 3.73) | 70 (61 - 78) | 6.3 (5.4 - 7.3) | 41.1 (37.8 - 44.4) | 2.2 (2.0 - 2.3) |
| 73 (69 - 86) | Stomach problems & Urinary problems | 1.33 (1.23 - 1.45) | 3.32 (3.03 - 3.62) | 76 (66 - 85) | 7.0 (5.9 - 8.2) | 36.5 (33.2 - 39.8) | 2.6 (2.4 - 2.8) |
| 74 (71 - 88) | Arthritis & Urinary problems | 1.30 (1.20 - 1.41) | 3.24 (2.96 - 3.52) | 72 (64 - 80) | 7.0 (5.9 - 8.2) | 36.8 (33.3 - 40.4) | 2.4 (2.3 - 2.6) |
| 75 (70 - 90) | Asthma/COPD & Thyroid disorder | 1.30 (1.18 - 1.42) | 3.23 (2.90 - 3.56) | 61 (52 - 69) | 6.8 (5.7 - 7.9) | 34.3 (30.9 - 37.8) | 1.9 (1.8 - 2.0) |
| 78 (71 - 90) | Depression/Anxiety & Thyroid disorder | 1.29 (1.18 - 1.40) | 3.20 (2.91 - 3.49) | 56 (49 - 64) | 6.8 (5.5 - 8.2) | 31.9 (28.5 - 35.3) | 1.8 (1.7 - 2.0) |
| 80 (71 - 91) | Musculoskeletal & Cancer | 1.28 (1.16 - 1.41) | 3.19 (2.86 - 3.51) | 79 (68 - 90) | 7.3 (6.3 - 8.2) | 29.9 (26.4 - 33.4) | 1.8 (1.7 - 1.9) |
| 82 (72 - 91) | Thyroid disorder & Stomach problems | 1.26 (1.15 - 1.37) | 3.14 (2.85 - 3.42) | 64 (56 - 73) | 7.0 (5.8 - 8.2) | 31.7 (28.3 - 35.2) | 2.3 (2.1 - 2.4) |
| 86 (73 - 98) | Stomach problems & Cardiovascular disease | 1.23 (1.12 - 1.34) | 3.06 (2.77 - 3.34) | 85 (74 - 95) | 6.8 (5.7 - 7.9) | 39.5 (36.2 - 42.9) | 3.0 (2.8 - 3.2) |
| 87 (74 - 98) | Urinary problems & Cardiovascular disease | 1.22 (1.13 - 1.33) | 3.05 (2.78 - 3.32) | 83 (73 - 93) | 7.5 (6.0 - 9.0) | 40.7 (36.8 - 44.6) | 2.9 (2.8 - 3.1) |
| 88 (74 - 98) | Diabetes & Depression/Anxiety | 1.22 (1.12 - 1.33) | 3.04 (2.76 - 3.32) | 71 (61 - 81) | 5.7 (4.3 - 7.2) | 46.9 (43.2 - 50.6) | 1.4 (1.3 - 1.5) |
| 89 (74 - 100) | Arthritis & Cardiovascular disease | 1.20 (1.09 - 1.31) | 2.99 (2.70 - 3.28) | 77 (67 - 88) | 6.2 (5.3 - 7.1) | 44.0 (39.9 - 48.1) | 2.7 (2.6 - 2.9) |
| 91 (79 - 102) | Asthma/COPD & Cardiovascular disease | 1.18 (1.08 - 1.28) | 2.93 (2.65 - 3.21) | 81 (71 - 92) | 5.9 (4.9 - 6.8) | 50.7 (46.7 - 54.7) | 2.5 (2.3 - 2.7) |
| 97 (84 - 106) | Thyroid disorder & Urinary problems | 1.13 (1.03 - 1.24) | 2.81 (2.54 - 3.09) | 59 (50 - 68) | 7.2 (5.9 - 8.6) | 30.3 (26.6 - 34.0) | 2.0 (1.8 - 2.1) |
| 101 (91 - 109) | Depression/Anxiety & Urinary problems | 1.09 (0.99 - 1.19) | 2.71 (2.45 - 2.98) | 58 (51 - 66) | 5.9 (4.7 - 7.1) | 37.7 (34.0 - 41.4) | 1.8 (1.7 - 1.9) |
| 107 (99 - 116) | Asthma/COPD & Urinary problems | 1.02 (0.94 - 1.11) | 2.55 (2.32 - 2.79) | 59 (52 - 66) | 5.4 (4.5 - 6.4) | 42.5 (38.6 - 46.4) | 1.8 (1.6 - 1.9) |
| 114 (104 - 133) | Thyroid disorder & Cardiovascular disease | 0.96 (0.87 - 1.06) | 2.39 (2.14 - 2.65) | 59 (50 - 68) | 5.6 (4.6 - 6.5) | 37.6 (32.7 - 42.4) | 2.1 (1.9 - 2.3) |
| 115 (105 - 134) | Hypertension & Stroke | 0.95 (0.86 - 1.05) | 2.37 (2.12 - 2.61) | 59 (50 - 68) | 4.1 (3.3 - 4.9) | 46.7 (42.9 - 50.5) | 2.9 (2.7 - 3.1) |

| | | | | | | | |
|---|---|---|---|---|---|---|---|
| 124 (109 - 138) | Hypertension & Heart Failure | 0.92 (0.84 - 1.01) | 2.30 (2.06 - 2.53) | 65 (57 - 73) | 4.9 (3.9 - 5.8) | 43.0 (38.5 - 47.6) | 2.8 (2.6 - 3.0) |
| 129 (115 - 146) | Thyroid disorder & Cancer | 0.87 (0.79 - 0.96) | 2.17 (1.96 - 2.39) | 47 (39 - 55) | 5.1 (4.3 - 6.0) | 27.0 (22.8 - 31.2) | 2.3 (2.2 - 2.5) |
| 132 (114 - 148) | Diabetes & Cancer | 0.87 (0.78 - 0.96) | 2.16 (1.93 - 2.40) | 59 (48 - 70) | 4.7 (3.8 - 5.6) | 33.8 (29.7 - 37.8) | 1.8 (1.6 - 1.9) |
| 134 (115 - 150) | Cardiovascular disease & Cancer | 0.86 (0.77 - 0.96) | 2.14 (1.90 - 2.38) | 63 (53 - 74) | 4.8 (4.0 - 5.6) | 36.1 (32.0 - 40.2) | 3.3 (3.0 - 3.5) |
| 135 (116 - 148) | Urinary problems & Cancer | 0.86 (0.78 - 0.95) | 2.14 (1.91 - 2.36) | 57 (49 - 66) | 5.3 (4.3 - 6.2) | 26.1 (22.0 - 30.1) | 2.5 (2.3 - 2.7) |
| 139 (125 - 154) | Arthritis & Cancer | 0.83 (0.76 - 0.91) | 2.07 (1.87 - 2.27) | 49 (42 - 55) | 4.8 (4.0 - 5.7) | 31.0 (27.1 - 34.9) | 2.3 (2.1 - 2.5) |
| 141 (126 - 156) | Stomach problems & Cancer | 0.82 (0.73 - 0.91) | 2.04 (1.81 - 2.26) | 54 (44 - 65) | 4.7 (3.9 - 5.6) | 28.9 (24.6 - 33.2) | 2.3 (2.1 - 2.5) |
| 144 (126 - 157) | Hyperlipidemia & Stroke | 0.80 (0.72 - 0.89) | 2.00 (1.78 - 2.22) | 49 (41 - 56) | 3.4 (2.7 - 4.1) | 44.8 (40.5 - 49.1) | 3.3 (3.1 - 3.6) |
| 151 (138 - 168) | Depression/Anxiety & Cardiovascular disease | 0.77 (0.69 - 0.85) | 1.91 (1.71 - 2.11) | 48 (42 - 55) | 3.6 (2.9 - 4.2) | 44.1 (39.4 - 48.7) | 1.6 (1.5 - 1.8) |
| 166 (149 - 180) | Hyperlipidemia & Heart Failure | 0.70 (0.63 - 0.78) | 1.75 (1.57 - 1.93) | 51 (44 - 59) | 3.9 (3.0 - 4.8) | 40.2 (35.4 - 45.1) | 2.9 (2.7 - 3.2) |
| 170 (154 - 186) | Asthma/COPD & Cancer | 0.69 (0.62 - 0.76) | 1.71 (1.52 - 1.90) | 42 (35 - 49) | 3.8 (3.1 - 4.5) | 42.0 (36.9 - 47.1) | 1.9 (1.7 - 2.0) |
| 186 (168 - 212) | Depression/Anxiety & Cancer | 0.62 (0.54 - 0.70) | 1.54 (1.33 - 1.74) | 40 (31 - 49) | 3.5 (2.7 - 4.4) | 29.9 (24.7 - 35.0) | 1.5 (1.3 - 1.7) |
| 192 (172 - 214) | Hypertension & Colon problems | 0.60 (0.53 - 0.68) | 1.50 (1.30 - 1.69) | 29 (25 - 34) | 3.4 (2.7 - 4.1) | 33.8 (28.3 - 39.3) | 1.6 (1.4 - 1.8) |
| 203 (181 - 223) | Cardiovascular disease & Heart Failure | 0.57 (0.50 - 0.65) | 1.42 (1.23 - 1.61) | 48 (39 - 56) | 3.0 (2.4 - 3.6) | 46.4 (41.2 - 51.7) | 7.4 (6.8 - 8.2) |
| 216 (198 - 240) | Musculoskeletal & Colon problems | 0.51 (0.45 - 0.59) | 1.28 (1.11 - 1.45) | 28 (22 - 33) | 2.7 (2.1 - 3.3) | 40.9 (35.2 - 46.7) | 2.3 (2.1 - 2.6) |
| 217 (201 - 240) | Hyperlipidemia & Colon problems | 0.51 (0.45 - 0.58) | 1.28 (1.11 - 1.45) | 26 (21 - 30) | 2.7 (2.1 - 3.3) | 31.3 (26.1 - 36.5) | 1.9 (1.7 - 2.1) |
| **Triad (3-way) multimorbidity combinations** | | | | | | | |
| 17 (16 - 18) | Hypertension & Hyperlipidemia & Diabetes | 4.62 (4.39 - 4.86) | 11.51 (10.82 - 12.21) | 216 (199 - 234) | 18.4 (16.6 - 20.2) | 34.3 (32.4 - 36.3) | 9.5 (9.3 - 9.8) |
| 29 (27 - 30) | Hypertension & Hyperlipidemia & Musculoskeletal | 3.16 (3.00 - 3.33) | 7.87 (7.39 - 8.35) | 160 (146 - 175) | 15.3 (13.0 - 17.6) | 34.4 (32.2 - 36.6) | 4.7 (4.5 - 4.8) |
| 33 (33 - 35) | Hypertension & Hyperlipidemia & Cardiovascular disease | 2.74 (2.57 - 2.91) | 6.82 (6.36 - 7.28) | 157 (142 - 172) | 12.3 (11.0 - 13.6) | 36.6 (34.0 - 39.2) | 10.0 (9.6 - 10.4) |
| 35 (33 - 39) | Hypertension & Hyperlipidemia & Arthritis | 2.59 (2.41 - 2.77) | 6.45 (5.97 - 6.92) | 124 (112 - 137) | 10.9 (9.6 - 12.1) | 37.9 (35.4 - 40.3) | 7.5 (7.2 - 7.8) |
| 38 (35 - 42) | Hypertension & Hyperlipidemia & Stomach problems | 2.44 (2.28 - 2.61) | 6.08 (5.62 - 6.53) | 124 (112 - 137) | 11.0 (9.7 - 12.2) | 33.2 (30.9 - 35.5) | 7.4 (7.1 - 7.7) |
| 43 (40 - 44) | Hypertension & Hyperlipidemia & Urinary problems | 2.23 (2.08 - 2.38) | 5.55 (5.15 - 5.96) | 119 (107 - 131) | 10.7 (9.5 - 12.0) | 34.2 (31.4 - 37.0) | 6.6 (6.3 - 6.9) |
| 46 (45 - 48) | Hypertension & Hyperlipidemia & Thyroid disorder | 1.99 (1.86 - 2.13) | 4.97 (4.61 - 5.33) | 87 (77 - 97) | 10.3 (8.3 - 12.4) | 29.0 (26.1 - 31.8) | 5.4 (5.1 - 5.6) |
| 48 (45 - 48) | Hypertension & Hyperlipidemia & Asthma/COPD | 1.94 (1.81 - 2.08) | 4.83 (4.45 - 5.22) | 99 (89 - 110) | 8.0 (7.0 - 9.0) | 42.8 (39.8 - 45.8) | 5.2 (4.9 - 5.4) |
| 50 (49 - 53) | Hypertension & Musculoskeletal & Arthritis | 1.76 (1.64 - 1.89) | 4.38 (4.04 - 4.73) | 94 (83 - 104) | 8.4 (7.3 - 9.6) | 41.2 (38.2 - 44.3) | 6.4 (6.0 - 6.7) |
| 52 (49 - 57) | Hypertension & Musculoskeletal & Diabetes | 1.69 (1.58 - 1.82) | 4.22 (3.90 - 4.54) | 99 (88 - 109) | 7.9 (6.8 - 8.9) | 44.1 (41.2 - 47.0) | 4.4 (4.2 - 4.7) |

| | | | | | | | |
|---|---|---|---|---|---|---|---|
| 56 (49 - 61) | Hypertension & Hyperlipidemia & Depression/Anxiety | 1.62 (1.49 - 1.77) | 4.04 (3.68 - 4.41) | 80 (70 - 90) | 7.2 (6.0 - 8.4) | 38.3 (35.0 - 41.6) | 4.1 (3.8 - 4.3) |
| 66 (58 - 72) | Hypertension & Hyperlipidemia & Cancer | 1.47 (1.35 - 1.59) | 3.66 (3.34 - 3.98) | 84 (72 - 95) | 7.3 (6.3 - 8.3) | 26.6 (23.7 - 29.5) | 6.4 (6.1 - 6.8) |
| 67 (59 - 72) | Hyperlipidemia & Musculoskeletal & Diabetes | 1.47 (1.36 - 1.58) | 3.65 (3.36 - 3.95) | 88 (77 - 98) | 7.0 (6.0 - 8.1) | 43.2 (39.9 - 46.4) | 5.3 (5.0 - 5.6) |
| 68 (58 - 72) | Hypertension & Musculoskeletal & Stomach problems | 1.46 (1.35 - 1.58) | 3.65 (3.33 - 3.96) | 82 (73 - 92) | 7.6 (6.5 - 8.8) | 40.0 (36.5 - 43.6) | 5.6 (5.3 - 5.9) |
| 69 (61 - 73) | Hypertension & Diabetes & Cardiovascular disease | 1.42 (1.30 - 1.54) | 3.53 (3.22 - 3.85) | 104 (91 - 116) | 6.8 (5.8 - 7.7) | 45.3 (41.7 - 48.9) | 9.4 (8.9 - 9.9) |
| 70 (61 - 73) | Hypertension & Diabetes & Arthritis | 1.42 (1.30 - 1.54) | 3.53 (3.22 - 3.83) | 83 (73 - 93) | 6.3 (5.4 - 7.3) | 47.2 (43.8 - 50.6) | 7.3 (6.9 - 7.7) |
| 71 (62 - 79) | Hyperlipidemia & Musculoskeletal & Arthritis | 1.38 (1.28 - 1.49) | 3.44 (3.16 - 3.73) | 77 (68 - 86) | 7.0 (6.1 - 8.0) | 38.7 (35.4 - 42.0) | 6.8 (6.4 - 7.2) |
| 76 (70 - 91) | Hypertension & Diabetes & Stomach problems | 1.29 (1.17 - 1.42) | 3.22 (2.90 - 3.53) | 84 (73 - 95) | 5.9 (4.9 - 6.8) | 44.6 (41.4 - 47.9) | 6.9 (6.5 - 7.3) |
| 81 (71 - 91) | Hyperlipidemia & Diabetes & Cardiovascular disease | 1.27 (1.17 - 1.39) | 3.17 (2.89 - 3.46) | 93 (81 - 105) | 6.2 (5.3 - 7.1) | 44.7 (40.9 - 48.4) | 11.4 (10.8 - 12.0) |
| 83 (73 - 91) | Hypertension & Diabetes & Urinary problems | 1.26 (1.15 - 1.37) | 3.13 (2.85 - 3.41) | 84 (74 - 95) | 6.3 (5.4 - 7.2) | 43.3 (39.5 - 47.1) | 6.6 (6.3 - 7.0) |
| 84 (73 - 98) | Hyperlipidemia & Diabetes & Arthritis | 1.23 (1.12 - 1.35) | 3.06 (2.77 - 3.35) | 73 (63 - 82) | 5.5 (4.5 - 6.4) | 45.7 (42.3 - 49.1) | 8.6 (8.1 - 9.1) |
| 85 (74 - 95) | Hypertension & Musculoskeletal & Asthma/COPD | 1.23 (1.13 - 1.33) | 3.06 (2.79 - 3.33) | 68 (60 - 77) | 6.1 (5.1 - 7.1) | 47.9 (44.2 - 51.6) | 4.2 (3.9 - 4.5) |
| 92 (82 - 104) | Hypertension & Arthritis & Stomach problems | 1.15 (1.04 - 1.27) | 2.87 (2.58 - 3.15) | 63 (54 - 72) | 5.5 (4.6 - 6.5) | 41.2 (37.6 - 44.8) | 8.5 (8.0 - 9.0) |
| 93 (84 - 104) | Hypertension & Musculoskeletal & Cardiovascular disease | 1.15 (1.06 - 1.25) | 2.86 (2.61 - 3.12) | 79 (69 - 88) | 5.6 (4.8 - 6.3) | 42.3 (38.7 - 45.8) | 5.4 (5.0 - 5.7) |
| 94 (84 - 106) | Hyperlipidemia & Diabetes & Stomach problems | 1.14 (1.04 - 1.26) | 2.85 (2.57 - 3.12) | 74 (62 - 86) | 5.2 (4.3 - 6.1) | 42.6 (39.1 - 46.1) | 8.4 (7.9 - 8.9) |
| 95 (85 - 104) | Hypertension & Musculoskeletal & Urinary problems | 1.13 (1.05 - 1.23) | 2.83 (2.59 - 3.07) | 70 (62 - 79) | 6.1 (5.3 - 7.0) | 39.6 (36.0 - 43.2) | 4.3 (4.0 - 4.6) |
| 96 (88 - 104) | Hyperlipidemia & Musculoskeletal & Stomach problems | 1.13 (1.04 - 1.22) | 2.82 (2.58 - 3.06) | 64 (56 - 71) | 6.1 (5.2 - 7.1) | 37.6 (34.2 - 40.9) | 6.1 (5.7 - 6.4) |
| 98 (84 - 107) | Hypertension & Musculoskeletal & Depression/Anxiety | 1.13 (1.02 - 1.24) | 2.81 (2.52 - 3.09) | 59 (51 - 66) | 5.3 (4.3 - 6.3) | 44.6 (40.4 - 48.8) | 3.7 (3.5 - 4.0) |
| 100 (89 - 108) | Hyperlipidemia & Diabetes & Urinary problems | 1.10 (1.00 - 1.20) | 2.73 (2.46 - 2.99) | 75 (65 - 85) | 5.5 (4.7 - 6.4) | 41.7 (37.5 - 45.8) | 8.0 (7.5 - 8.5) |
| 102 (91 - 110) | Hypertension & Diabetes & Asthma/COPD | 1.08 (0.98 - 1.19) | 2.69 (2.42 - 2.96) | 67 (58 - 76) | 4.6 (4.0 - 5.3) | 53.2 (49.3 - 57.1) | 5.1 (4.8 - 5.5) |
| 105 (96 - 114) | Hypertension & Musculoskeletal & Thyroid disorder | 1.04 (0.96 - 1.13) | 2.60 (2.38 - 2.82) | 59 (51 - 67) | 7.3 (5.4 - 9.2) | 33.0 (29.4 - 36.7) | 3.5 (3.3 - 3.8) |
| 106 (92 - 116) | Hypertension & Diabetes & Thyroid disorder | 1.04 (0.94 - 1.15) | 2.59 (2.32 - 2.87) | 58 (49 - 67) | 5.5 (4.4 - 6.6) | 36.8 (33.0 - 40.5) | 5.0 (4.7 - 5.3) |

| | | | | | | | |
|---|---|---|---|---|---|---|---|
| 108 (99 - 124) | Hypertension & Asthma/COPD & Arthritis | 1.02 (0.92 - 1.12) | 2.53 (2.26 - 2.80) | 55 (47 - 63) | 4.2 (3.5 - 4.8) | 47.8 (43.7 - 52.0) | 6.8 (6.3 - 7.2) |
| 109 (101 - 124) | Hyperlipidemia & Musculoskeletal & Cardiovascular disease | 1.00 (0.91 - 1.09) | 2.49 (2.25 - 2.72) | 66 (57 - 76) | 4.8 (4.2 - 5.4) | 39.5 (35.5 - 43.4) | 6.3 (5.9 - 6.7) |
| 110 (102 - 126) | Hypertension & Asthma/COPD & Stomach problems | 0.98 (0.89 - 1.08) | 2.44 (2.18 - 2.71) | 58 (50 - 66) | 4.5 (3.7 - 5.3) | 44.6 (40.3 - 48.9) | 6.9 (6.5 - 7.4) |
| 111 (103 - 126) | Hyperlipidemia & Diabetes & Asthma/COPD | 0.98 (0.89 - 1.08) | 2.44 (2.18 - 2.69) | 60 (51 - 68) | 4.1 (3.5 - 4.7) | 52.3 (47.9 - 56.8) | 6.2 (5.8 - 6.6) |
| 112 (102 - 128) | Hyperlipidemia & Diabetes & Thyroid disorder | 0.97 (0.87 - 1.08) | 2.42 (2.16 - 2.69) | 55 (46 - 64) | 5.1 (4.1 - 6.2) | 36.0 (32.1 - 39.9) | 6.4 (6.0 - 6.8) |
| 116 (105 - 137) | Hypertension & Arthritis & Cardiovascular disease | 0.95 (0.85 - 1.05) | 2.36 (2.10 - 2.61) | 62 (53 - 70) | 4.8 (4.0 - 5.6) | 47.8 (43.2 - 52.4) | 8.3 (7.8 - 8.9) |
| 117 (107 - 137) | Hypertension & Stomach problems & Cardiovascular disease | 0.93 (0.85 - 1.03) | 2.33 (2.09 - 2.56) | 66 (57 - 76) | 5.2 (4.1 - 6.2) | 41.8 (37.8 - 45.7) | 8.7 (8.1 - 9.3) |
| 117 (107 - 137) | Hyperlipidemia & Arthritis & Stomach problems | 0.93 (0.85 - 1.03) | 2.33 (2.09 - 2.57) | 53 (46 - 60) | 5.1 (4.2 - 6.0) | 40.0 (36.0 - 43.9) | 9.5 (8.9 - 10.2) |
| 119 (108 - 137) | Hyperlipidemia & Musculoskeletal & Urinary problems | 0.93 (0.85 - 1.02) | 2.32 (2.10 - 2.55) | 60 (51 - 68) | 5.1 (4.4 - 5.9) | 37.2 (33.2 - 41.3) | 4.7 (4.4 - 5.1) |
| 121 (109 - 137) | Hyperlipidemia & Musculoskeletal & Thyroid disorder | 0.93 (0.85 - 1.01) | 2.32 (2.10 - 2.53) | 50 (43 - 58) | 6.1 (4.3 - 7.9) | 30.6 (26.9 - 34.3) | 4.3 (4.0 - 4.6) |
| 122 (108 - 138) | Hypertension & Stomach problems & Urinary problems | 0.93 (0.84 - 1.02) | 2.31 (2.08 - 2.54) | 54 (47 - 61) | 4.8 (3.9 - 5.6) | 39.3 (35.3 - 43.4) | 7.2 (6.7 - 7.7) |
| 123 (109 - 137) | Hyperlipidemia & Musculoskeletal & Asthma/COPD | 0.92 (0.84 - 1.01) | 2.30 (2.09 - 2.51) | 55 (48 - 62) | 4.3 (3.7 - 4.8) | 48.7 (44.4 - 52.9) | 4.4 (4.1 - 4.7) |
| 125 (109 - 139) | Hypertension & Urinary problems & Cardiovascular disease | 0.91 (0.83 - 1.00) | 2.27 (2.04 - 2.49) | 65 (56 - 73) | 5.4 (4.4 - 6.3) | 44.4 (40.0 - 48.9) | 8.5 (7.9 - 9.1) |
| 126 (112 - 143) | Hypertension & Arthritis & Urinary problems | 0.89 (0.81 - 0.97) | 2.21 (1.99 - 2.43) | 52 (46 - 59) | 4.4 (3.8 - 5.1) | 42.7 (38.4 - 47.0) | 6.5 (6.1 - 7.0) |
| 127 (111 - 145) | Hypertension & Diabetes & Depression/Anxiety | 0.89 (0.80 - 0.98) | 2.21 (1.97 - 2.45) | 54 (46 - 63) | 4.1 (2.8 - 5.4) | 46.9 (42.7 - 51.1) | 3.9 (3.7 - 4.2) |
| 130 (114 - 147) | Hyperlipidemia & Stomach problems & Cardiovascular disease | 0.87 (0.79 - 0.96) | 2.17 (1.95 - 2.40) | 58 (50 - 65) | 4.5 (3.7 - 5.3) | 39.0 (35.0 - 43.1) | 11.1 (10.3 - 11.9) |
| 131 (112 - 148) | Hypertension & Depression/Anxiety & Stomach problems | 0.87 (0.78 - 0.97) | 2.17 (1.91 - 2.42) | 51 (42 - 59) | 4.7 (3.1 - 6.3) | 44.2 (39.9 - 48.6) | 5.7 (5.3 - 6.1) |
| 132 (114 - 148) | Hypertension & Asthma/COPD & Cardiovascular disease | 0.87 (0.78 - 0.96) | 2.16 (1.93 - 2.40) | 63 (54 - 71) | 4.4 (3.6 - 5.2) | 53.7 (49.2 - 58.2) | 7.2 (6.7 - 7.8) |
| 135 (116 - 149) | Hyperlipidemia & Musculoskeletal & Depression/Anxiety | 0.86 (0.77 - 0.95) | 2.14 (1.92 - 2.36) | 45 (39 - 51) | 4.3 (3.4 - 5.3) | 44.5 (40.3 - 48.7) | 3.9 (3.6 - 4.2) |
| 137 (116 - 150) | Hypertension & Arthritis & Thyroid disorder | 0.86 (0.77 - 0.95) | 2.13 (1.91 - 2.35) | 44 (38 - 50) | 4.8 (3.9 - 5.8) | 38.3 (33.9 - 42.8) | 5.5 (5.1 - 6.0) |
| 138 (117 - 154) | Hyperlipidemia & Urinary problems & Cardiovascular disease | 0.84 (0.76 - 0.94) | 2.10 (1.87 - 2.34) | 59 (51 - 68) | 4.7 (3.9 - 5.5) | 42.1 (37.3 - 47.0) | 10.4 (9.7 - 11.2) |
| 140 (125 - 156) | Hyperlipidemia & Arthritis & Cardiovascular disease | 0.82 (0.73 - 0.92) | 2.05 (1.82 - 2.29) | 53 (45 - 62) | 4.0 (3.3 - 4.8) | 45.2 (40.2 - 50.3) | 9.9 (9.2 - 10.6) |

| | | | | | | | |
|---|---|---|---|---|---|---|---|
| 142 (126 - 156) | Hypertension & Depression/Anxiety & Arthritis | 0.82 (0.73 - 0.91) | 2.03 (1.81 - 2.26) | 47 (40 - 54) | 3.8 (3.1 - 4.6) | 46.2 (41.8 - 50.6) | 5.3 (4.9 - 5.7) |
| 145 (126 - 157) | Hyperlipidemia & Asthma/COPD & Arthritis | 0.80 (0.72 - 0.89) | 2.00 (1.78 - 2.22) | 47 (40 - 54) | 3.3 (2.8 - 3.8) | 46.5 (42.3 - 50.7) | 7.3 (6.8 - 7.8) |
| 146 (128 - 158) | Musculoskeletal & Arthritis & Stomach problems | 0.80 (0.72 - 0.88) | 1.99 (1.78 - 2.19) | 49 (41 - 56) | 4.6 (3.7 - 5.5) | 43.2 (38.9 - 47.6) | 10.2 (9.5 - 11.0) |
| 147 (128 - 163) | Hyperlipidemia & Diabetes & Depression/Anxiety | 0.79 (0.71 - 0.87) | 1.97 (1.75 - 2.19) | 49 (41 - 58) | 3.8 (2.9 - 4.8) | 46.4 (42.1 - 50.8) | 4.8 (4.5 - 5.2) |
| 149 (138 - 165) | Hyperlipidemia & Asthma/COPD & Stomach problems | 0.78 (0.71 - 0.85) | 1.93 (1.74 - 2.12) | 48 (41 - 55) | 3.6 (3.1 - 4.2) | 43.3 (39.0 - 47.5) | 7.5 (7.0 - 8.1) |
| 153 (138 - 170) | Hyperlipidemia & Stomach problems & Urinary problems | 0.76 (0.69 - 0.85) | 1.90 (1.70 - 2.11) | 46 (39 - 53) | 4.1 (3.3 - 4.8) | 38.3 (34.0 - 42.7) | 8.2 (7.6 - 8.9) |
| 155 (139 - 171) | Musculoskeletal & Asthma/COPD & Arthritis | 0.75 (0.67 - 0.84) | 1.87 (1.66 - 2.08) | 46 (39 - 53) | 3.6 (2.9 - 4.3) | 49.4 (44.9 - 54.0) | 8.5 (7.9 - 9.2) |
| 156 (141 - 174) | Hypertension & Thyroid disorder & Stomach problems | 0.74 (0.67 - 0.82) | 1.85 (1.65 - 2.04) | 42 (35 - 49) | 4.7 (3.8 - 5.7) | 34.8 (30.3 - 39.3) | 5.3 (4.9 - 5.7) |
| 157 (144 - 178) | Hyperlipidemia & Arthritis & Urinary problems | 0.73 (0.65 - 0.81) | 1.81 (1.61 - 2.01) | 44 (37 - 51) | 3.9 (3.1 - 4.7) | 41.6 (36.8 - 46.4) | 7.3 (6.7 - 7.9) |
| 158 (144 - 180) | Musculoskeletal & Depression/Anxiety & Arthritis | 0.72 (0.64 - 0.81) | 1.80 (1.58 - 2.01) | 41 (35 - 47) | 3.3 (2.7 - 3.9) | 42.8 (38.0 - 47.6) | 7.8 (7.2 - 8.4) |
| 159 (144 - 180) | Hypertension & Musculoskeletal & Cancer | 0.72 (0.64 - 0.80) | 1.79 (1.59 - 2.00) | 46 (39 - 53) | 4.1 (3.4 - 4.8) | 35.5 (31.2 - 39.7) | 4.0 (3.6 - 4.3) |
| 159 (144 - 180) | Hypertension & Depression/Anxiety & Asthma/COPD | 0.72 (0.64 - 0.81) | 1.79 (1.58 - 2.01) | 40 (34 - 45) | 2.8 (2.3 - 3.4) | 49.4 (44.3 - 54.4) | 4.3 (4.0 - 4.7) |
| 162 (146 - 180) | Musculoskeletal & Diabetes & Arthritis | 0.72 (0.64 - 0.80) | 1.79 (1.59 - 1.99) | 48 (41 - 56) | 3.7 (3.0 - 4.5) | 51.9 (47.0 - 56.7) | 6.2 (5.8 - 6.7) |
| 163 (146 - 180) | Hyperlipidemia & Asthma/COPD & Cardiovascular disease | 0.71 (0.64 - 0.80) | 1.78 (1.57 - 1.99) | 51 (43 - 59) | 3.5 (2.8 - 4.1) | 51.1 (46.1 - 56.0) | 8.1 (7.5 - 8.8) |
| 164 (146 - 180) | Hyperlipidemia & Arthritis & Thyroid disorder | 0.71 (0.64 - 0.80) | 1.78 (1.58 - 1.98) | 36 (31 - 42) | 4.0 (3.2 - 4.8) | 37.1 (32.0 - 42.2) | 6.2 (5.7 - 6.8) |
| 166 (149 - 180) | Hypertension & Asthma/COPD & Thyroid disorder | 0.70 (0.63 - 0.78) | 1.75 (1.56 - 1.95) | 36 (31 - 42) | 4.1 (3.2 - 4.9) | 37.7 (33.4 - 42.0) | 4.2 (3.9 - 4.6) |
| 172 (155 - 192) | Hypertension & Thyroid disorder & Cardiovascular disease | 0.67 (0.60 - 0.75) | 1.68 (1.48 - 1.87) | 45 (37 - 53) | 3.9 (3.1 - 4.7) | 41.9 (36.5 - 47.3) | 5.8 (5.3 - 6.3) |
| 173 (156 - 192) | Hyperlipidemia & Depression/Anxiety & Arthritis | 0.67 (0.60 - 0.75) | 1.67 (1.48 - 1.86) | 38 (32 - 44) | 3.5 (2.8 - 4.2) | 44.1 (39.7 - 48.5) | 5.7 (5.3 - 6.2) |
| 174 (155 - 195) | Musculoskeletal & Asthma/COPD & Stomach problems | 0.67 (0.59 - 0.75) | 1.67 (1.46 - 1.87) | 43 (35 - 50) | 3.8 (2.9 - 4.8) | 47.8 (42.7 - 52.9) | 7.9 (7.3 - 8.5) |
| 175 (157 - 197) | Hypertension & Hyperlipidemia & Stroke | 0.66 (0.59 - 0.74) | 1.64 (1.45 - 1.84) | 41 (34 - 48) | 2.8 (2.1 - 3.4) | 48.0 (43.3 - 52.7) | 10.6 (9.8 - 11.4) |
| 176 (157 - 200) | Hypertension & Thyroid disorder & Urinary problems | 0.66 (0.58 - 0.74) | 1.64 (1.44 - 1.83) | 39 (32 - 47) | 4.2 (3.3 - 5.2) | 35.9 (30.9 - 40.8) | 4.7 (4.3 - 5.1) |
| 177 (157 - 201) | Hypertension & Diabetes & Cancer | 0.65 (0.58 - 0.74) | 1.63 (1.43 - 1.83) | 46 (37 - 55) | 3.6 (2.8 - 4.5) | 34.2 (29.7 - 38.6) | 5.0 (4.6 - 5.5) |

| | | | | | | | |
|---|---|---|---|---|---|---|---|
| 178 (158 - 197) | Musculoskeletal & Arthritis & Thyroid disorder | 0.65 (0.59 - 0.72) | 1.63 (1.45 - 1.80) | 36 (30 - 42) | 4.2 (3.5 - 5.0) | 35.0 (30.5 - 39.5) | 7.0 (6.5 - 7.7) |
| 179 (157 - 201) | Hyperlipidemia & Depression/Anxiety & Stomach problems | 0.65 (0.58 - 0.73) | 1.62 (1.43 - 1.82) | 39 (31 - 46) | 3.4 (2.7 - 4.1) | 43.9 (39.5 - 48.3) | 5.9 (5.4 - 6.4) |
| 180 (157 - 201) | Hyperlipidemia & Thyroid disorder & Stomach problems | 0.65 (0.58 - 0.73) | 1.61 (1.43 - 1.80) | 36 (30 - 41) | 3.8 (3.0 - 4.7) | 33.1 (28.3 - 37.9) | 6.4 (5.9 - 7.0) |
| 182 (168 - 204) | Musculoskeletal & Depression/Anxiety & Asthma/COPD | 0.63 (0.56 - 0.70) | 1.56 (1.38 - 1.75) | 37 (31 - 43) | 2.8 (2.2 - 3.4) | 50.6 (45.1 - 56.2) | 6.6 (6.1 - 7.2) |
| 184 (169 - 204) | Hypertension & Asthma/COPD & Urinary problems | 0.62 (0.56 - 0.69) | 1.55 (1.38 - 1.73) | 40 (34 - 45) | 3.2 (2.6 - 3.8) | 48.2 (43.3 - 53.1) | 4.3 (4.0 - 4.7) |
| 184 (166 - 209) | Hypertension & Cardiovascular disease & Cancer | 0.62 (0.55 - 0.70) | 1.55 (1.36 - 1.75) | 45 (37 - 53) | 3.5 (2.8 - 4.2) | 37.0 (32.1 - 41.9) | 8.8 (8.0 - 9.5) |
| 189 (169 - 214) | Musculoskeletal & Depression/Anxiety & Stomach problems | 0.61 (0.53 - 0.69) | 1.52 (1.32 - 1.71) | 37 (30 - 44) | 3.0 (2.3 - 3.6) | 42.0 (36.5 - 47.5) | 6.9 (6.4 - 7.6) |
| 191 (174 - 212) | Musculoskeletal & Arthritis & Urinary problems | 0.60 (0.54 - 0.67) | 1.50 (1.34 - 1.66) | 38 (32 - 43) | 3.9 (3.0 - 4.7) | 41.8 (36.9 - 46.7) | 7.5 (6.9 - 8.1) |
| 195 (172 - 214) | Hyperlipidemia & Thyroid disorder & Cardiovascular disease | 0.59 (0.52 - 0.67) | 1.48 (1.29 - 1.67) | 38 (30 - 45) | 3.2 (2.5 - 3.8) | 39.4 (33.2 - 45.6) | 7.0 (6.4 - 7.6) |
| 196 (175 - 214) | Musculoskeletal & Diabetes & Stomach problems | 0.59 (0.53 - 0.66) | 1.47 (1.30 - 1.64) | 41 (35 - 48) | 3.1 (2.5 - 3.8) | 49.8 (45.2 - 54.5) | 5.6 (5.1 - 6.1) |
| 196 (175 - 214) | Hyperlipidemia & Asthma/COPD & Thyroid disorder | 0.59 (0.52 - 0.66) | 1.47 (1.28 - 1.65) | 32 (27 - 38) | 3.2 (2.6 - 3.8) | 39.3 (34.4 - 44.2) | 4.8 (4.4 - 5.3) |
| 199 (175 - 214) | Hyperlipidemia & Musculoskeletal & Cancer | 0.58 (0.52 - 0.66) | 1.46 (1.27 - 1.64) | 39 (31 - 46) | 3.4 (2.7 - 4.0) | 32.5 (27.7 - 37.3) | 4.4 (4.0 - 4.9) |
| 200 (179 - 214) | Hypertension & Hyperlipidemia & Heart Failure | 0.58 (0.52 - 0.65) | 1.45 (1.29 - 1.62) | 46 (39 - 53) | 3.3 (2.5 - 4.1) | 41.4 (36.0 - 46.8) | 9.1 (8.3 - 9.9) |
| 202 (181 - 220) | Hypertension & Depression/Anxiety & Thyroid disorder | 0.57 (0.51 - 0.64) | 1.43 (1.26 - 1.59) | 30 (25 - 36) | 3.1 (2.3 - 3.8) | 38.9 (34.0 - 43.8) | 3.4 (3.1 - 3.8) |
| 203 (179 - 224) | Hyperlipidemia & Thyroid disorder & Urinary problems | 0.57 (0.50 - 0.65) | 1.42 (1.23 - 1.61) | 33 (26 - 40) | 3.3 (2.6 - 4.1) | 34.8 (29.3 - 40.4) | 5.5 (5.1 - 6.1) |
| 205 (181 - 230) | Hyperlipidemia & Depression/Anxiety & Asthma/COPD | 0.56 (0.49 - 0.63) | 1.39 (1.21 - 1.57) | 31 (26 - 36) | 2.5 (1.9 - 3.0) | 51.3 (45.4 - 57.3) | 4.6 (4.2 - 5.0) |
| 205 (182 - 229) | Hypertension & Urinary problems & Cancer | 0.56 (0.49 - 0.63) | 1.39 (1.21 - 1.57) | 37 (31 - 44) | 3.2 (2.6 - 3.9) | 28.8 (23.8 - 33.9) | 6.1 (5.6 - 6.7) |
| 207 (181 - 232) | Hypertension & Depression/Anxiety & Urinary problems | 0.56 (0.49 - 0.63) | 1.39 (1.20 - 1.57) | 32 (27 - 37) | 2.6 (2.1 - 3.2) | 43.8 (38.7 - 48.9) | 3.8 (3.5 - 4.2) |
| 209 (184 - 233) | Hyperlipidemia & Diabetes & Cancer | 0.55 (0.48 - 0.63) | 1.37 (1.19 - 1.55) | 42 (32 - 51) | 3.0 (2.3 - 3.6) | 35.1 (30.3 - 39.9) | 6.0 (5.5 - 6.6) |
| 210 (186 - 232) | Hypertension & Stomach problems & Cancer | 0.55 (0.48 - 0.62) | 1.37 (1.20 - 1.54) | 36 (29 - 44) | 3.2 (2.4 - 3.9) | 30.9 (25.9 - 35.9) | 6.0 (5.4 - 6.6) |

| | | | | | | | |
|---|---|---|---|---|---|---|---|
| 211 (186 - 233) | Musculoskeletal & Diabetes & Asthma/COPD | 0.55 (0.48 - 0.62) | 1.36 (1.19 - 1.54) | 37 (30 - 43) | 2.5 (2.0 - 2.9) | 57.0 (52.1 - 61.8) | 4.5 (4.1 - 4.9) |
| 212 (186 - 234) | Hyperlipidemia & Cardiovascular disease & Cancer | 0.54 (0.48 - 0.62) | 1.36 (1.18 - 1.54) | 41 (33 - 49) | 2.9 (2.3 - 3.5) | 34.6 (29.6 - 39.6) | 10.8 (9.8 - 11.8) |
| 213 (190 - 233) | Hypertension & Arthritis & Cancer | 0.54 (0.48 - 0.61) | 1.35 (1.19 - 1.50) | 34 (28 - 39) | 3.1 (2.4 - 3.8) | 35.8 (30.8 - 40.9) | 5.7 (5.2 - 6.3) |
| 214 (191 - 233) | Musculoskeletal & Diabetes & Urinary problems | 0.54 (0.48 - 0.60) | 1.34 (1.19 - 1.49) | 42 (35 - 50) | 3.0 (2.4 - 3.5) | 50.6 (45.3 - 55.8) | 5.0 (4.5 - 5.4) |
| 215 (198 - 240) | Musculoskeletal & Stomach problems & Urinary problems | 0.52 (0.45 - 0.59) | 1.29 (1.12 - 1.45) | 34 (29 - 40) | 3.1 (2.4 - 3.9) | 41.9 (36.5 - 47.4) | 7.1 (6.5 - 7.7) |
| 217 (200 - 240) | Hyperlipidemia & Depression/Anxiety & Thyroid disorder | 0.51 (0.45 - 0.58) | 1.28 (1.11 - 1.45) | 24 (20 - 29) | 2.7 (2.1 - 3.4) | 39.1 (33.4 - 44.8) | 4.1 (3.7 - 4.5) |
| 217 (199 - 240) | Hypertension & Depression/Anxiety & Cardiovascular disease | 0.51 (0.45 - 0.58) | 1.28 (1.10 - 1.45) | 35 (29 - 41) | 2.4 (1.8 - 3.0) | 47.4 (41.7 - 53.2) | 4.3 (3.9 - 4.8) |
| 220 (202 - 240) | Musculoskeletal & Thyroid disorder & Stomach problems | 0.51 (0.45 - 0.58) | 1.27 (1.11 - 1.43) | 33 (27 - 38) | 3.6 (2.8 - 4.4) | 37.0 (31.8 - 42.3) | 6.4 (5.8 - 7.0) |
| 221 (202 - 240) | Hyperlipidemia & Asthma/COPD & Urinary problems | 0.51 (0.45 - 0.58) | 1.27 (1.10 - 1.43) | 34 (29 - 40) | 2.9 (2.2 - 3.5) | 48.9 (43.4 - 54.5) | 4.8 (4.4 - 5.3) |
| 224 (205 - 240) | Musculoskeletal & Arthritis & Cardiovascular disease | 0.50 (0.44 - 0.57) | 1.24 (1.08 - 1.41) | 34 (28 - 39) | 2.7 (2.1 - 3.2) | 41.7 (36.2 - 47.2) | 7.6 (6.9 - 8.4) |
| 225 (205 - 240) | Musculoskeletal & Diabetes & Cardiovascular disease | 0.50 (0.44 - 0.56) | 1.24 (1.08 - 1.40) | 40 (33 - 47) | 2.5 (2.0 - 3.0) | 48.9 (43.1 - 54.7) | 5.7 (5.1 - 6.2) |
| 229 (205 - 240) | Diabetes & Arthritis & Stomach problems | 0.50 (0.44 - 0.56) | 1.24 (1.08 - 1.39) | 36 (30 - 42) | 2.8 (2.1 - 3.6) | 51.3 (45.9 - 56.7) | 8.9 (8.1 - 9.7) |
| 230 (205 - 240) | Asthma/COPD & Arthritis & Stomach problems | 0.49 (0.43 - 0.56) | 1.22 (1.06 - 1.39) | 30 (25 - 35) | 2.4 (1.7 - 3.0) | 44.4 (39.3 - 49.6) | 11.8 (10.8 - 12.9) |
| 231 (205 - 240) | Hyperlipidemia & Urinary problems & Cancer | 0.49 (0.43 - 0.56) | 1.22 (1.05 - 1.39) | 36 (29 - 42) | 3.0 (2.3 - 3.7) | 27.4 (22.5 - 32.3) | 7.6 (6.9 - 8.4) |
| 232 (209 - 240) | Musculoskeletal & Asthma/COPD & Thyroid disorder | 0.49 (0.43 - 0.56) | 1.22 (1.05 - 1.38) | 28 (23 - 33) | 3.3 (2.6 - 4.1) | 38.7 (33.3 - 44.1) | 4.9 (4.4 - 5.4) |
| 235 (212 - 240) | Hyperlipidemia & Depression/Anxiety & Cardiovascular disease | 0.48 (0.42 - 0.55) | 1.19 (1.02 - 1.35) | 30 (25 - 36) | 2.2 (1.6 - 2.7) | 44.7 (38.8 - 50.7) | 5.3 (4.8 - 5.9) |
| 238 (214 - 240) | Diabetes & Asthma/COPD & Arthritis | 0.47 (0.41 - 0.54) | 1.18 (1.02 - 1.34) | 33 (27 - 40) | 2.1 (1.7 - 2.5) | 58.6 (53.1 - 64.1) | 7.5 (6.9 - 8.3) |
| 239 (215 - 240) | Hypertension & Thyroid disorder & Cancer | 0.47 (0.41 - 0.53) | 1.17 (1.02 - 1.32) | 27 (22 - 33) | 2.9 (2.2 - 3.6) | 30.2 (24.7 - 35.7) | 5.0 (4.5 - 5.5) |
| 240 (215 - 240) | Musculoskeletal & Stomach problems & Cardiovascular disease | 0.47 (0.41 - 0.53) | 1.16 (1.01 - 1.31) | 36 (30 - 43) | 2.5 (2.0 - 3.0) | 46.1 (40.7 - 51.5) | 7.8 (7.1 - 8.6) |
| **Tetrad (4-way) multimorbidity combinations** | | | | | | | |
| 90 (74 - 99) | Hypertension & Hyperlipidemia & Musculoskeletal & Diabetes | 1.20 (1.10 - 1.30) | 2.98 (2.72 - 3.25) | 75 (65 - 84) | 5.8 (4.8 - 6.8) | 43.8 (40.2 - 47.4) | 16.5 (15.6 - 17.5) |
| 99 (89 - 107) | Hypertension & Hyperlipidemia & Diabetes & Cardiovascular disease | 1.11 (1.02 - 1.21) | 2.77 (2.51 - 3.03) | 83 (72 - 94) | 5.2 (4.4 - 6.0) | 44.9 (40.9 - 49.0) | 38.6 (36.4 - 40.9) |

| | | | | | | | |
|---|---|---|---|---|---|---|---|
| 103 (91 - 111) | Hypertension & Hyperlipidemia & Diabetes & Arthritis | 1.07 (0.97 - 1.18) | 2.68 (2.41 - 2.94) | 65 (56 - 74) | 4.7 (3.9 - 5.6) | 46.3 (42.6 - 50.0) | 28.9 (27.3 - 30.6) |
| 104 (92 - 111) | Hypertension & Hyperlipidemia & Musculoskeletal & Arthritis | 1.06 (0.97 - 1.16) | 2.64 (2.40 - 2.89) | 62 (53 - 70) | 5.3 (4.5 - 6.1) | 42.2 (38.5 - 46.0) | 20.3 (19.1 - 21.5) |
| 113 (104 - 130) | Hypertension & Hyperlipidemia & Diabetes & Stomach problems | 0.97 (0.87 - 1.07) | 2.41 (2.16 - 2.66) | 64 (54 - 74) | 4.5 (3.6 - 5.3) | 43.3 (39.5 - 47.1) | 27.6 (25.9 - 29.3) |
| 119 (107 - 138) | Hypertension & Hyperlipidemia & Diabetes & Urinary problems | 0.93 (0.84 - 1.03) | 2.32 (2.08 - 2.57) | 64 (55 - 73) | 4.7 (3.9 - 5.5) | 44.2 (39.6 - 48.8) | 26.2 (24.6 - 27.9) |
| 128 (115 - 145) | Hypertension & Hyperlipidemia & Musculoskeletal & Stomach problems | 0.87 (0.80 - 0.96) | 2.18 (1.97 - 2.39) | 52 (45 - 59) | 4.8 (4.0 - 5.7) | 40.1 (36.1 - 44.2) | 18.2 (17.1 - 19.5) |
| 143 (126 - 156) | Hypertension & Hyperlipidemia & Musculoskeletal & Cardiovascular disease | 0.81 (0.74 - 0.89) | 2.02 (1.81 - 2.23) | 55 (47 - 64) | 3.8 (3.2 - 4.4) | 41.9 (37.5 - 46.2) | 19.9 (18.6 - 21.4) |
| 148 (131 - 167) | Hypertension & Hyperlipidemia & Diabetes & Asthma/COPD | 0.78 (0.70 - 0.87) | 1.95 (1.73 - 2.17) | 50 (42 - 59) | 3.3 (2.8 - 3.9) | 54.5 (49.7 - 59.3) | 19.3 (18.0 - 20.7) |
| 152 (134 - 171) | Hypertension & Hyperlipidemia & Diabetes & Thyroid disorder | 0.77 (0.68 - 0.86) | 1.91 (1.68 - 2.14) | 45 (37 - 53) | 4.2 (3.2 - 5.2) | 37.2 (32.9 - 41.5) | 19.4 (18.1 - 20.8) |
| 154 (138 - 171) | Hypertension & Hyperlipidemia & Arthritis & Stomach problems | 0.76 (0.68 - 0.85) | 1.89 (1.68 - 2.10) | 44 (37 - 51) | 4.0 (3.1 - 4.8) | 42.0 (37.5 - 46.5) | 29.8 (27.8 - 32.0) |
| 159 (146 - 180) | Hypertension & Hyperlipidemia & Stomach problems & Cardiovascular disease | 0.72 (0.65 - 0.80) | 1.79 (1.60 - 1.99) | 49 (42 - 56) | 3.7 (3.0 - 4.4) | 40.5 (35.9 - 45.0) | 35.4 (32.9 - 38.2) |
| 165 (148 - 180) | Hypertension & Hyperlipidemia & Musculoskeletal & Urinary problems | 0.71 (0.64 - 0.79) | 1.78 (1.59 - 1.96) | 49 (42 - 57) | 4.0 (3.3 - 4.7) | 41.5 (36.9 - 46.0) | 14.1 (13.1 - 15.3) |
| 168 (148 - 186) | Hypertension & Hyperlipidemia & Arthritis & Cardiovascular disease | 0.70 (0.62 - 0.79) | 1.74 (1.52 - 1.95) | 47 (39 - 55) | 3.4 (2.7 - 4.1) | 47.7 (42.4 - 53.0) | 32.5 (30.1 - 35.0) |
| 169 (153 - 185) | Hypertension & Hyperlipidemia & Musculoskeletal & Asthma/COPD | 0.69 (0.62 - 0.76) | 1.72 (1.53 - 1.90) | 43 (37 - 50) | 3.2 (2.7 - 3.7) | 51.1 (46.1 - 56.1) | 12.7 (11.8 - 13.7) |
| 171 (153 - 187) | Hypertension & Hyperlipidemia & Urinary problems & Cardiovascular disease | 0.68 (0.61 - 0.76) | 1.70 (1.50 - 1.90) | 50 (42 - 58) | 3.8 (3.1 - 4.6) | 44.4 (39.2 - 49.7) | 32.8 (30.4 - 35.5) |
| 181 (166 - 204) | Hypertension & Hyperlipidemia & Asthma/COPD & Arthritis | 0.63 (0.56 - 0.71) | 1.57 (1.38 - 1.76) | 38 (31 - 44) | 2.4 (2.0 - 2.9) | 49.0 (44.5 - 53.4) | 22.6 (21.0 - 24.4) |
| 182 (166 - 206) | Hypertension & Hyperlipidemia & Diabetes & Depression/Anxiety | 0.63 (0.56 - 0.70) | 1.56 (1.37 - 1.75) | 41 (33 - 49) | 3.0 (2.1 - 3.9) | 47.4 (42.5 - 52.3) | 14.9 (13.8 - 16.2) |
| 187 (169 - 210) | Hypertension & Hyperlipidemia & Stomach problems & Urinary problems | 0.62 (0.55 - 0.69) | 1.53 (1.35 - 1.71) | 38 (32 - 44) | 3.3 (2.6 - 4.0) | 40.2 (35.3 - 45.1) | 25.8 (23.8 - 27.9) |
| 188 (172 - 210) | Hypertension & Hyperlipidemia & Musculoskeletal & Thyroid disorder | 0.61 (0.55 - 0.68) | 1.52 (1.36 - 1.68) | 36 (30 - 42) | 4.5 (2.8 - 6.3) | 33.3 (28.6 - 37.9) | 11.1 (10.2 - 12.1) |
| 190 (172 - 214) | Hypertension & Hyperlipidemia & Asthma/COPD & Cardiovascular disease | 0.60 (0.53 - 0.68) | 1.50 (1.32 - 1.69) | 45 (37 - 53) | 2.8 (2.3 - 3.4) | 51.3 (45.9 - 56.6) | 26.7 (24.6 - 28.9) |

| | | | | | | | |
|---|---|---|---|---|---|---|---|
| 193 (172 - 214) | Hypertension & Hyperlipidemia & Musculoskeletal & Depression/Anxiety | 0.60 (0.53 - 0.68) | 1.49 (1.31 - 1.67) | 34 (28 - 39) | 3.1 (2.2 - 4.0) | 48.1 (42.8 - 53.5) | 10.5 (9.7 - 11.5) |
| 194 (175 - 214) | Hypertension & Hyperlipidemia & Asthma/COPD & Stomach problems | 0.60 (0.53 - 0.66) | 1.48 (1.31 - 1.65) | 39 (33 - 45) | 2.8 (2.3 - 3.3) | 46.2 (41.1 - 51.2) | 22.7 (21.0 - 24.6) |
| 198 (175 - 214) | Hypertension & Hyperlipidemia & Arthritis & Urinary problems | 0.59 (0.52 - 0.66) | 1.46 (1.28 - 1.64) | 37 (31 - 43) | 2.9 (2.4 - 3.5) | 44.8 (39.3 - 50.2) | 22.6 (20.9 - 24.6) |
| 201 (179 - 216) | Hypertension & Musculoskeletal & Diabetes & Arthritis | 0.58 (0.51 - 0.65) | 1.44 (1.27 - 1.62) | 40 (33 - 47) | 3.1 (2.4 - 3.8) | 53.4 (47.9 - 58.9) | 19.7 (18.2 - 21.3) |
| 207 (184 - 229) | Hypertension & Musculoskeletal & Arthritis & Stomach problems | 0.56 (0.49 - 0.63) | 1.39 (1.22 - 1.55) | 36 (29 - 42) | 3.2 (2.4 - 3.9) | 47.2 (42.0 - 52.4) | 27.6 (25.4 - 29.9) |
| 221 (201 - 240) | Hypertension & Hyperlipidemia & Arthritis & Thyroid disorder | 0.51 (0.44 - 0.58) | 1.27 (1.10 - 1.43) | 28 (23 - 33) | 2.9 (2.1 - 3.6) | 43.8 (37.8 - 49.8) | 17.6 (16.1 - 19.2) |
| 223 (205 - 240) | Hyperlipidemia & Musculoskeletal & Diabetes & Arthritis | 0.50 (0.44 - 0.57) | 1.25 (1.09 - 1.41) | 36 (29 - 42) | 2.8 (2.1 - 3.5) | 51.2 (45.6 - 56.8) | 22.9 (21.0 - 25.0) |
| 225 (205 - 240) | Hypertension & Hyperlipidemia & Depression/Anxiety & Stomach problems | 0.50 (0.43 - 0.57) | 1.24 (1.07 - 1.41) | 32 (25 - 40) | 2.6 (2.0 - 3.3) | 46.8 (41.6 - 52.0) | 17.5 (16.0 - 19.1) |
| 225 (205 - 240) | Hypertension & Hyperlipidemia & Depression/Anxiety & Arthritis | 0.50 (0.44 - 0.57) | 1.24 (1.07 - 1.40) | 31 (25 - 36) | 2.6 (1.9 - 3.2) | 47.0 (41.4 - 52.6) | 16.6 (15.2 - 18.2) |
| 225 (205 - 240) | Hypertension & Musculoskeletal & Diabetes & Stomach problems | 0.50 (0.44 - 0.56) | 1.24 (1.08 - 1.40) | 36 (30 - 42) | 2.8 (2.1 - 3.4) | 50.5 (45.3 - 55.7) | 18.1 (16.6 - 19.8) |
| 233 (209 - 240) | Hypertension & Musculoskeletal & Asthma/COPD & Arthritis | 0.48 (0.42 - 0.55) | 1.21 (1.04 - 1.37) | 30 (25 - 36) | 2.4 (1.8 - 2.9) | 52.5 (46.8 - 58.3) | 21.5 (19.7 - 23.5) |
| 234 (212 - 240) | Hypertension & Hyperlipidemia & Thyroid disorder & Stomach problems | 0.48 (0.42 - 0.54) | 1.19 (1.04 - 1.35) | 27 (22 - 32) | 3.1 (2.3 - 3.9) | 35.8 (30.2 - 41.4) | 18.2 (16.6 - 19.9) |
| 237 (211 - 240) | Hypertension & Hyperlipidemia & Diabetes & Cancer | 0.48 (0.41 - 0.55) | 1.19 (1.02 - 1.35) | 37 (28 - 45) | 2.6 (2.0 - 3.3) | 36.2 (30.8 - 41.5) | 19.5 (17.8 - 21.5) |

**Supplementary Figure 2: Highly prevalent specific combinations of multimorbidity and associated cost and perceived poor health, excluding hypertension and hyperlipidemia**

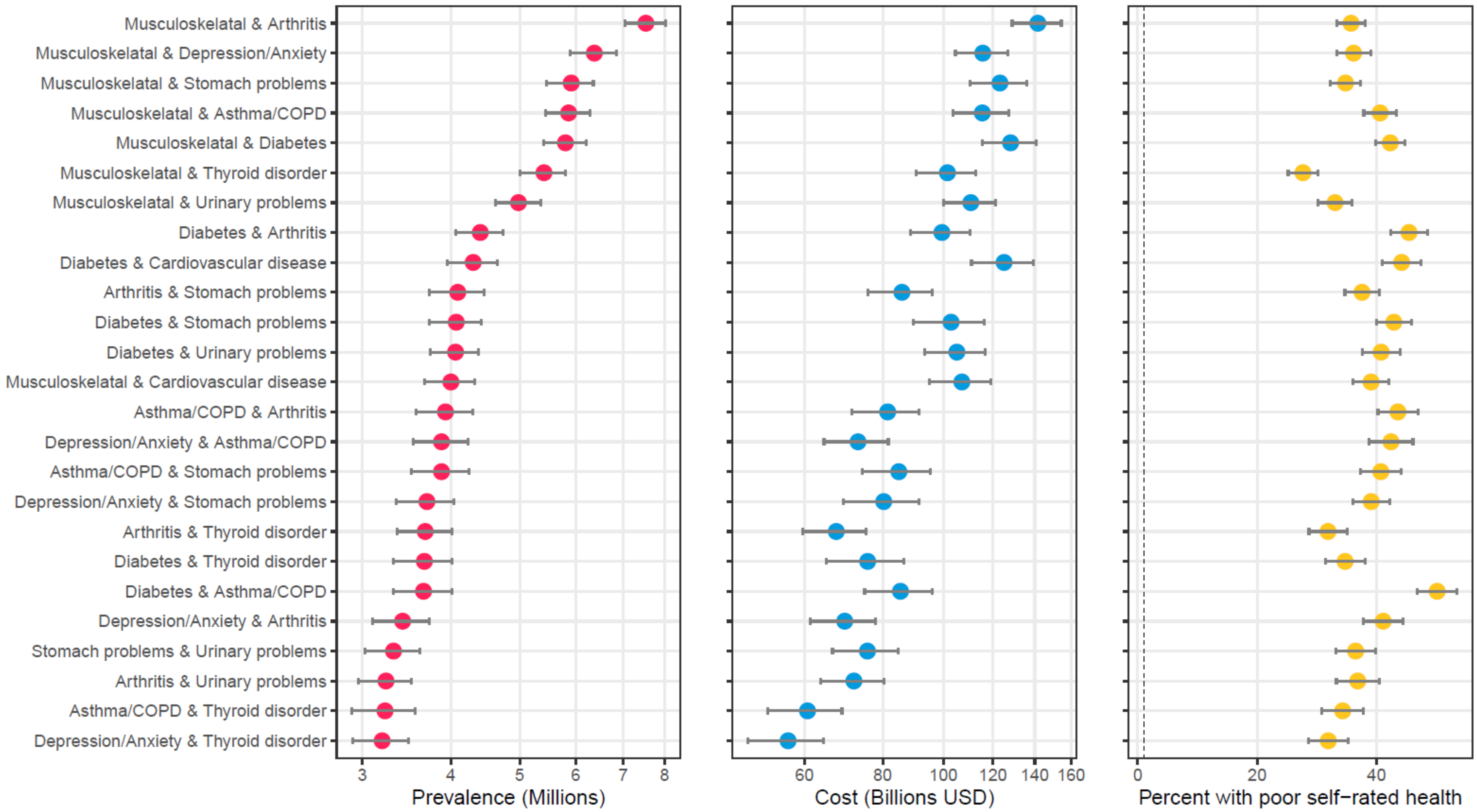

**Supplementary Figure 3: Specific multimorbidity combinations with highest average out-of-pocket expenditures among U.S. adults, 2016-2019**

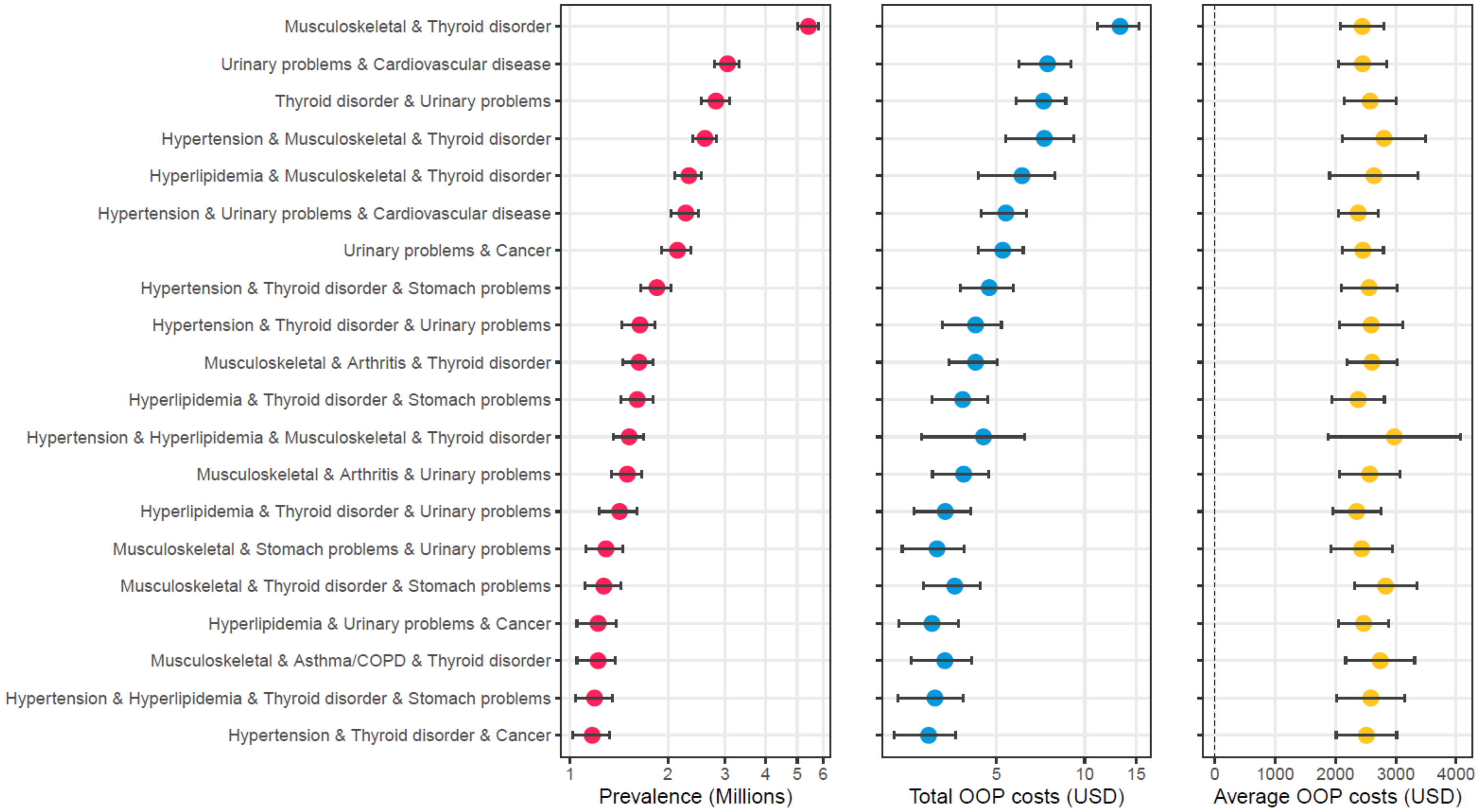
